# Quantifying Clinical Trial Diversity of FDA Novel Drug Approvals

**DOI:** 10.1101/2023.05.11.23289884

**Authors:** William E Fitzsimmons, Muhammed Y Idris, Priscilla Pemu

**Affiliations:** University of Illinois at Chicago Colleges of Pharmacy and Medicine, Chicago, IL, USA; Clinical Research Center, Morehouse School of Medicine, Atlanta, GA, USA; CARER Group, San Diego, CA, USA

**Keywords:** Clinical Trial Diversity, FDA approvals, Novel Drugs

## Abstract

**Background:** Health care inequity includes the lack of adequate representation of various populations in clinical trials. Government, academic and industry organizations have highlighted these issues and committed to actions to improve. In order to assess the current status and future success of these initiatives a quantitative objective measure to assess the state of clinical trial diversity is needed.

**Methods:** FDA review documents for all novel drug approvals from January 2022 through February 16, 2023 were assessed using a scorecard that considers diversity across different demographic subgroups including age (>65 yo), sex (female), race (Black and Asian) and ethnicity (Hispanic/Latino). The scorecard assigns each drug a letter grade, between A and F, for each subgroup (and overall) based on 1) the percent of each sub-population included in the trials and grades relative to the percent of the US population, 2) the number of participants from each subpopulation that received the novel new drug in the trials, 3) the incidence or prevalence of the disease/condition in each of the sub-populations.

**Results:** The FDA approved 43 novel new drugs for 44 indications (one drug was simultaneously approved for two indications). The three drugs with A Grades reflecting the best diversity in their registration trials were tapinarof (Vtama from Dermavant), daprodustat (Jesduvroq from GlaxoSmithKline) and eflapegrastim (Rolvedon from Spectrum Pharmaceuticals.) There was good representation of elderly and females with only two drugs receiving a D grade in either of these sub-populations. In contrast, Black and Hispanic representation was often inadequate with 4 drugs receiving F grades. There were 9 drugs (20%) where there were no Black participants receiving the novel new drug and an additional 14 approvals where there were <10 Black participants receiving the novel drug. The median number of Black participants receiving the investigational drug was 9. In the Hispanic/Latino population there were 2 approvals with no Hispanic participants receiving the novel drug and 14 approvals where there were < 10 Hispanic participants receiving the drug. The median number of Hispanic participants receiving the novel drug was 12.5.

**Conclusions:** This newly developed scorecard provides an objective quantitative approach to assess the current state of diversity in clinical trials supporting new drug approvals. Substantial improvement in racial and ethnic representation is needed. Meaningful change will require actions and cooperation amongst all stakeholders to address this multifaceted issue and will take commitment, perseverance, and appropriate incentives.

## Background

Despite the rapid pace of scientific discovery driven by technological innovation, efforts to translate those discoveries into solutions that address persistent health disparities have been considerably less effective. A particularly challenging problem is the fact that clinical trials, the seminal first step in delivery of new medicines and therapies, typically do not adequately include the diverse populations who live in our most underserved and marginalized communities. Because safety and effectiveness may vary in different populations, the lack of diversity in clinical trial enrollment compromises the health care that can be delivered to those who are excluded. Whereas this problem has been recognized for decades, progress has been slow in overcoming this critical deficiency.

Awareness and the “Calls for Action” for Clinical Trial Diversity took major steps forward in 2022 on the U.S. national front. The year was marked by reports from: The National Academy of Sciences, Engineering, and Medicine report on Improving Representation in Clinical Trials and Research: Building Research Equity for Women and Underrepresented which highlighted the lack of progress in increasing trial participation of racial and ethnic minority population groups and the subsequent impact on health disparities and the national costs and consequences; The Government Accountability Office on Practices to Facilitate Diversity of Patients in Cancer Clinical Trials which focused on the Federal actions that have been taken to facilitate diversity in cancer clinical trials and the best practices from 17 cancer centers with a history of enrolling diverse populations.^1,2^ The FDA published a Draft Guidance entitled “Diversity Plans to Improve Enrollment of Participants from Underrepresented Racial and Ethnic Populations in Clinical Trials Guidance for Industry” to provide recommendations to biopharmaceutical sponsors developing medical products for developing a Race and Ethnicity Diversity Plan to enroll representative numbers of racial and ethnic populations in the United States in clinical trials.^3^ Legislatively, numerous Bills were also introduced into the 117th Congress with measures to address diversity in clinical trials including the DEPICT Act (H.R. 6584), the CURES 2.0 Act (H.R. 6000), the DIVERSE Trials Act (H.R. 5030, S.2706), and the ENACT Act of 2021 (H.R. 3085, S.1548). Although these Bills were not passed, on December 29,2022 the President signed H.R.2617 - Consolidated Appropriations Act, 2023 into Law which included amendments to laws governing the FDA referred to as Food and Drug Omnibus Reform Act of 2022, or FDORA.^4^ These amendments included guidance, workshops, summary reports and requirements for diversity action plans for clinical studies. While all of these reports are indicators of positive momentum and a desire to improve clinical trial diversity, there is a lack of quantitative objective data and evidence to assess the state of clinical trial diversity and in turn evaluate novel new drug approvals.

## Methods

A methodology was developed to assess the clinical trial diversity in novel drug approvals by the U.S. as tabulated by calendar year in the FDA website.^5^ Starting in January 2022 through February 16, 2023, the FDA review for each novel drug, including new molecular entities and new therapeutic biological products, was evaluated as published in the FDA review documents.^6^ FDA reviews provided a breakdown of the demography for the pivotal registration trial(s) including age (≥65 yo), sex (female), race (Black or African, Asian) and ethnicity (Hispanic or Latino).

### Analyzing Clinical Trial Diversity

The demography for the pivotal registration trial(s) that demonstrated the safety and efficacy of the new drug were assessed.

### Percent in Clinical Trial

The percentage of the total population enrolled for each group was calculated across both the treatment and control groups and compared to the U.S. Census data for each group and graded as *below* (less than 70% of the census representation), *meets* (within 70-130%) or *exceeds* (>130% of the census representation).^7^ The 30% relative margin was chosen to reflect a significant deviation from the census data which encompasses a two sided 90% confidence interval around the population average (assuming a sample size of 300) and accounts for the challenges of enrolling at or above the census average percentage.

### Number Treated with New Drug

Next, the total number of each group that were treated with the new drug was tabulated for the pivotal trial(s). The main focus for this metric was to assess the ability to detect safety events in the group that were “very common” (>10%), or “common” (>1%) per EMA SmPC guidance.^8^ The “Rule of Three’s” was used to define whether there was sufficient exposure to detect a very common event (i.e. at least 30) or a common event (i.e. at least 300).^9^

### Incidence of Disease or Condition

Subsequently a literature search was performed to determine if the disease or condition for which the new drug was approved is seen in decreased, similar or increased incidence/prevalence in the group compared to others (e.g. <65 yo, male, white non-Hispanic).

These three metrics, percent enrolled in pivotal trials, number treated with novel drug, and incidence/prevalence in the group, were color coded in the scorecard and used to develop a grade (A-F) for each diversity group per the algorithm shown in the Supplemental material. The grades across the 5 groups were averaged (A=4, B=3, C=2, D=1, F=0) and an overall diversity grade for the novel drug was assigned based on the scoring shown in the supplemental material.

## Results

From January, 2022 through February 16, 2023, the FDA approved 43 novel new drugs for 44 indications (one drug was simultaneously approved for two indications). An analysis of the grades for these 43 drugs is summarized in Table 1 and the scorecard for each of these 44 indications is included in the Supplemental material. The three drugs with A Grades reflecting the best diversity in their registration trials were tapinarof (Vtama from Dermavant), Jesduvroq (daprodustat from GlaxoSmithKline), and eflapegrastim (Rolvedon from Spectrum Pharmaceuticals) (Figure 1). In general, there was good representation of elderly and females with only two drugs receiving a D grade in either of these groups. In contrast, Black and Hispanic representation was markedly inadequate with 4 drugs receiving F grades.

**Table 1.** Novel Drug Approvals in the US from January 2022 through February 16, 2023; 43 drugs and 44 indications

|  | Overall | Elderly* | Female* | Black | Asian | Hispanic |
| --- | --- | --- | --- | --- | --- | --- |
| Grade | Grade<br>(N)** |  |  |  |  |  |
| A | 3 | 11 | 13 | 0 | 13 | 5 |
| B | 15 | 17 | 16 | 5 | 10 | 4 |
| C | 23 | 12 | 10 | 17 | 16 | 19 |
| D | 3 | 2 | 2 | 18 | 5 | 12 |
| F | 0 | 0 | 0 | 4 | 0 | 4 |

\* 2 elderly and 4 female drug reviews were excluded due to limited focus of the indications
\*\* Overall Grade combining all diversity groups

There were 9 drugs (20%; Table 2) where there were no Black participants receiving the novel new drug and an additional 14 approvals where there were <10 Black participants receiving the novel drug. Although several of the indications reflected in the 9 drugs in Table 2 have a lower incidence in Black compared to White racial groups, overall there were 11 (25%) drugs where the incidence/prevalence of the indication is increased in Blacks however the percent of Black participants in the pivotal trial(s) was less than 9.4% (i.e. <70% of the 13.4% of the U.S. census). The median number of Black participants receiving the investigational drug across all 44 indications was 9.

**Table 2.**
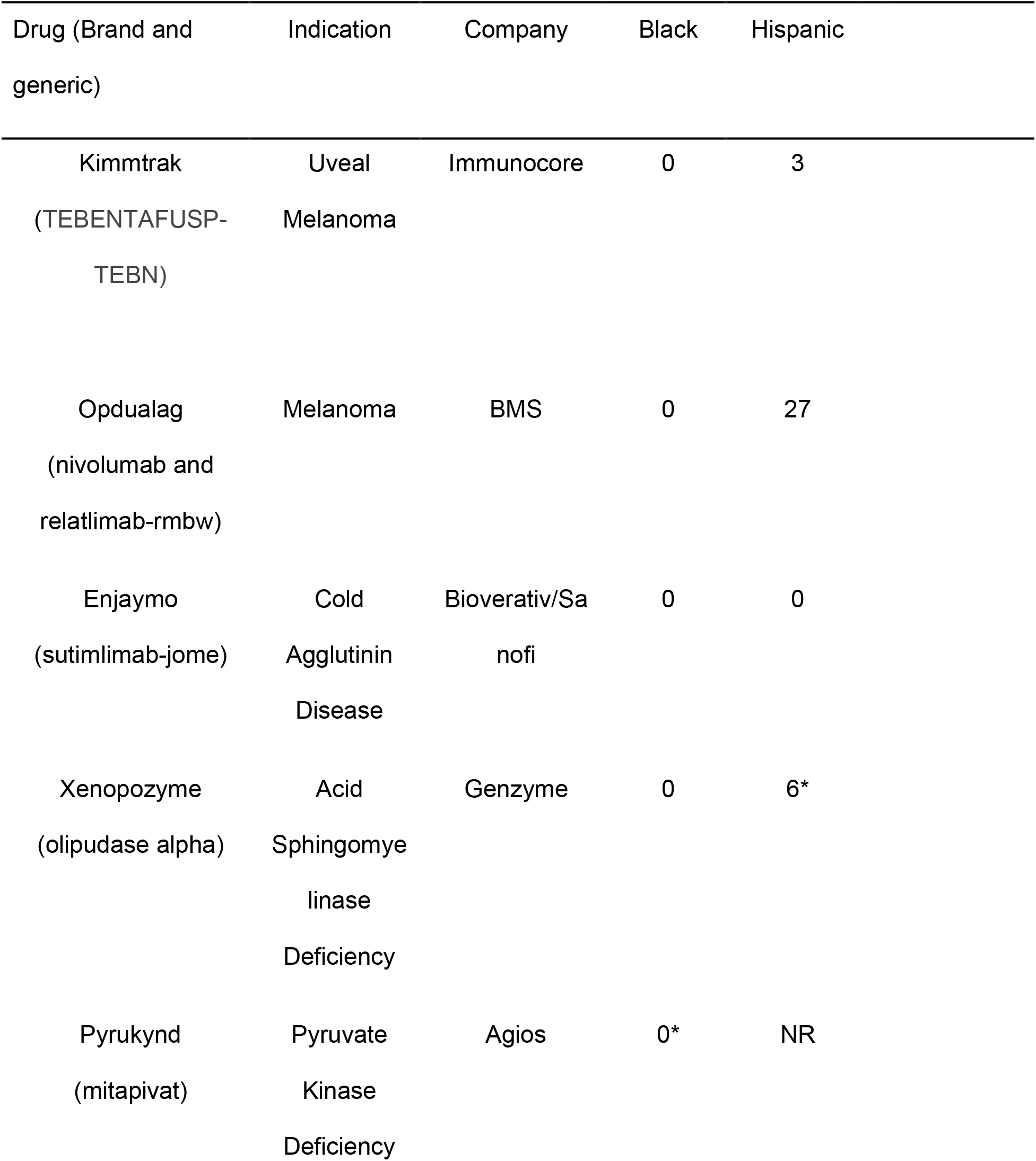
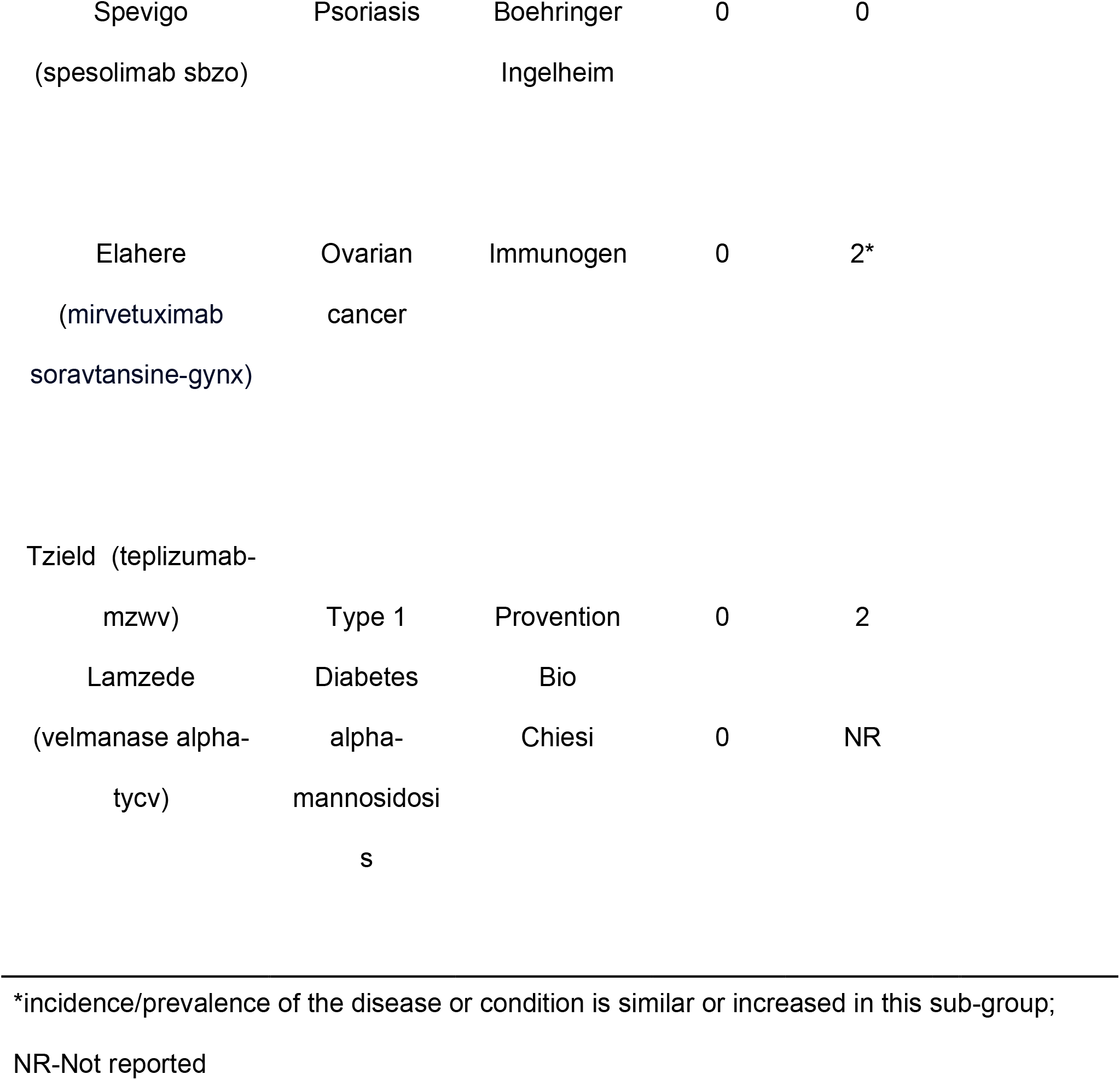
Novel Drug Approvals with 0 Black and/or Hispanic Study Participants receiving Novel Drug

In the Hispanic/Latino population there were 2 approvals with no Hispanic participants receiving the novel drug (shown in Table 2) and 14 approvals where there were < 10 Hispanic participants receiving the drug. The median number of Hispanic participants receiving the novel drug was 12.5. There were 3 drugs where the incidence/prevalence of the indication is increased in Hispanics but the percent of Hispanic participants in the pivotal trial(s) was less than 13% (i.e. <70% of the 18.5% of the U.S. census). Additionally, there were 8 approvals where the FDA did not report Hispanic representation.

To further evaluate the potential reasons for under-representation of U.S. racial and ethnic groups, the enrollment by country was assessed. Enrollment by geography was reported in 32 of the FDA drug reviews. The average enrollment from the U.S or U.S./Canada was 43% with a median enrollment of 33%.

## Conclusions

Based on this analysis of all recent novel drug approvals by the U.S. FDA, Phase 3 pivotal trials do not consistently represent the Black and Hispanic population impacted by these diseases and conditions in the U.S. This is in contrast with the FDA’s analysis of pivotal trials from 2015-2019 where they concluded that mean and yearly participation of Black participants was at or above US census data.^10^ In general, there is good representation of the elderly, females and Asian racial group.

This scorecard was developed to be objective, quantitative, oriented to the U.S. population, and focused on individual drugs/biologics different from previous ranking focusing on pharmaceutical companies.^11^ Limitations of the scorecard include: ordinal direction of the incidence/prevalence of the condition in the group (increased, similar or decreased); dependence on reporting in the FDA review documents, label and published literature; some countries (e.g. France) do not report race/ethnicity; does not include indigenous and Native American, Native Hawaian and Other Pacific Islanders; doesn’t account for gender and sexual identity.

Multidisciplinary efforts to enhance diversity are essential and we applaud the biopharmaceutical industry, federal government, and academic community for taking initial steps in the right direction.^12-15^ Continued and sustained efforts are essential. It is recommended that the FDA: 1) consider Post-Marketing Commitments as was done for mirvetuximab soravtansine-gynx (Elahere) for ovarian cancer in lieu of Post-marketing Requirement authority; 2) Standardize reporting of demographics in drug reviews. Ethnicity was not reported in several FDA reviews and there were differences in racial groups included in the data summaries. Biopharmaceutical Industry and organizations (PhRMA and BIO) should: 1) share best practices, including Diversity Action Plans, with transparency; 2) consider “onshoring” trials vs. offshoring as described by FDA Commissioner Califf; 3) incentivize Contract Research Organizations and study teams to enroll diverse populations 4) reassess limits on compensation to study participants. Legislators must 1) advance further legislation to incentivize industry (priority reviews or voucher, Diversity designations (similar to orphan designations); 2) advance aspects of the DEPICT, CURES 2.0, ENACT and DIVERSE Trials acts that were not included in FDORA. Investigators and clinical research centers, particularly historically black and minority service institutions, must be empowered to 1) advance the proposed clinical trial diversity scorecard to consider other racial groups (e.g. Indigenous and Native American, Native Hawaiian and Other Pacific Islanders) and underrepresented populations (e.g., sexual and gender minorities); 2) catalyze interactions between stakeholders and 3) continue sharing relevant practices to facilitate diverse enrollment and address barriers to enrollment.

Diverse representation is essential to ensure the safety and efficacy is evaluated in relevant groups before approval and commercialization.^16-17^ Meaningful change will require actions and cooperation amongst all stakeholders to address this multifaceted issue and will take commitment, perseverance, and appropriate incentives.

## Data Availability

All data produced are available online at https://www.accessdata.fda.gov/scripts/cder/daf/index.cfm

