## Supplemental Materials for "Quantifying Clinical Trial Diversity of FDA Novel Drug Approvals"

### CARER Clinical Trial Diversity Scorecard

shutterstock.com · 1514513114

- Includes new molecular entities and new therapeutic biological products that FDA CDER approved starting in 2022 https://www.fda.gov/drugs/new-drugs-fda-cders-new-molecular-entities-and-new-therapeutic-biological-products/novel-drug-approvals-2022
- Diversity metrics are focused on underrepresented populations defined by age, sex, race, and ethnicity
- Demographic data based on FDA Review documents published at and includes the populations from the pivotal registration trials and referenced at bottom of scorecard
- Percent of population includes subjects in the control arms
- TOTAL treated with new drug does not include control arms
- Literature supporting demography for each disease/condition referenced at bottom of scorecard
- U.S. population benchmarks from US Census Bureau https://www.census.gov/quickfacts/fact/table/US/PST045221

### CARER Clinical Trial Diversity Scorecard

shutterstock.com · 1514513114

- **Clinical Trial percent graded on +/- 30% relative difference from the U.S. population estimates (e.g. Black or African 13.4% X 0.3 = 4.0%; therefore similar to population estimates is 9.4%-17.4%).**

- **Grade for a given demographic subgroup is based on:**
  - **Qualitative assessment of Incidence/Prevalence of the disease and or condition in the demographic subgroup (references at the bottom of the scorecard)**
  - **% of the demographic subgroup in the study population. If data on a demographic group is not reported (NR) then grade is an F if disease is increased, D if disease is similar and C if disease is decreased in the demographic subgroup.**
  - **Total number of individuals in the demographic subgroup who have been treated with the new molecular entity. Note: N=30 is needed to detect a "very common" adverse reaction which occurs with an incidence of 10% or greater; N=300 is needed to detect a "common" adverse reaction which occur with an incidence of 1% or greater.**
  - **If the disease/condition is restricted only to males or females (e.g. prostate cancer in men, vulvovaginitis in women) the Sex category will not be graded and will be given an NA.**
  - **See table on the following slide for details on grading**

- **Overall Grade averages all the demographic subgroup grades as follows:**
  - **Each subgroup is graded from A to F with A=4 and F=0.**
  - **The total score for the grades are averaged across the total number of subgroups**
  - **If ANY of the subgroups have a D or F grade, then an overall Grade of A will not be assigned. If the average is in the A range, then a B will be assigned.**
  - **Overall Grade is defined as follows:**
    - **A= 3.25-4.0*(None of the subgroups have a D or F grade)**
    - **B= 2.5-3.25**
    - **C=1.75-2.5**
    - **D=1.0-1.75**
    - **F=<1.0**

| Incidence/Prevalence of the Disease or Condition in the demographic subgroup | % of the demographic subgroup in the clinical trials relative to U.S. Population | Grade if the number of individuals in the demographic subgroup treated with new Drug <30 | Grade if the number of individuals in the demographic subgroup treated with new Drug >30 | Grade if the number of individuals in the demographic subgroup treated with new Drug >300 |
|---|---|---|---|---|
| Increased | Above | C | B | A |
| Increased | Same | D | C | B |
| Increased | Below | F | D | C |
| Similar | Above | B | A | A |
| Similar | Same | C | B | A |
| Similar | Below | D | C | B |
| Decreased | Above | A | A | A |
| Decreased | Same | B | A | A |
| Decreased | Below | C | B | A |

### CARER Clinical Trial Diversity Scorecard

| Drug | |
|---|---|
| Tradename | **Quviviq** |
| Generic Name | **daridorexant** |
| Company | Idorsia |
| Date of FDA Approval | January 7, 2022 |
| Indication | treatment of adult patients with **insomnia** characterized by difficulties with sleep onset and/or sleep maintenance |

**OVERALL GRADE**

# B

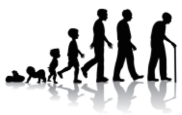

Age

| Age | % in U.S. Population | % in Clinical Trials | Number treated with new drug | Incidence of Disease or Condition | Grade |
|---|---|---|---|---|---|
| >/=65 years old | 16.5% | 39% | 484 | Increased | **A** |

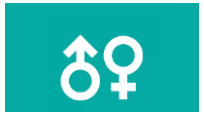

Sex

| Sex | % in U.S. Population | % in Clinical Trials | Number treated with new drug | Incidence of Disease or Condition | Grade |
|---|---|---|---|---|---|
| Female | 50.8% | 69% | 847 | Increased | **A** |

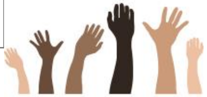

Race

| Race | % in U.S. Population | % in Clinical Trials | Number treated with new drug | Incidence of disease or condition | Grade |
|---|---|---|---|---|---|
| Black or African | 13.4% | 8% | 91 | Increased | **D** |
| Asian | 5.9% | 2% | 32 | Similar | **C** |

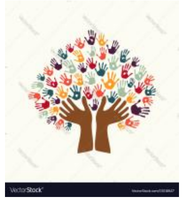

Ethnicity

| Ethnicity | % in U.S. Population | % in Clinical Trials | Number treated with new drug | Incidence of disease or condition | Grade |
|---|---|---|---|---|---|
| Hispanic or Latino | 18.5% | 10% | 126 | Similar | **C** |

**References: doi:10.3389/fpsyt.2020.577429; doi:10.5664/jcsm.7172; doi: 10.1016/j.socscimed.2016.02.012; doi:10.3390/brainsci9110306;**
**Three Phase 3 trials : ID-078A301 and ID078A302**
**https://www.accessdata.fda.gov/drugsatfda_docs/nda/2022/214985Orig1s000IntegratedR.pdf {page 36)**

### CARER Clinical Trial Diversity Scorecard

| Drug | |
|---|---|
| Tradename | **Cibinqo** |
| Generic Name | **abrocitinib** |
| Company | Pfizer |
| Date of FDA Approval | January 14, 2022 |
| Indication | treatment of adults with refractory, **moderate-to-severe atopic dermatitis** whose disease is not adequately controlled with other systemic drug products, including biologics, or when use of those therapies is inadvisable. |

#### OVERALL GRADE

# C

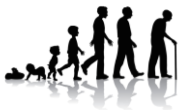

Age

| **Age** | **% in U.S. Population** | **% in Clinical Trials** | **Number treated with new drug** | **Incidence of Disease or Condition** | **Grade** |
|---|---|---|---|---|---|
| >/=65 years old | 16.5% | 5.5% | 62 | Increased | **D** |

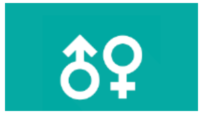

Sex

| **Sex** | **% in U.S. Population** | **% in Clinical Trials** | **Number treated with new drug** | **Incidence of Disease or Condition** | **Grade** |
|---|---|---|---|---|---|
| Female | 50.8% | 46.9% | 510 | Similar | **A** |

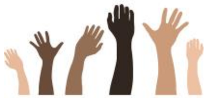

Race

| **Race** | **% in U.S. Population** | **% in Clinical Trials** | **Number treated with new drug** | **Incidence of disease or condition** | **Grade** |
|---|---|---|---|---|---|
| Black or African | 13.4% | 5.4% | 56 | Increased | **D** |
| Asian | 5.9% | 11.7% | 153 | Increased | **B** |

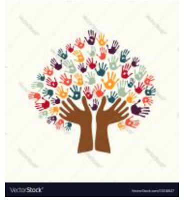

Ethnicity

| **Ethnicity** | **% in U.S. Population** | **% in Clinical Trials** | **Number treated with new drug** | **Incidence of disease or condition** | **Grade** |
|---|---|---|---|---|---|
| Hispanic or Latino | 18.5% | NR | NR | Similar | **D** |

**References: doi: 10.1111/exd.13514. PMID: 29457272. https://doi.org/10.1371/journal.pone.0258219 https://doi.org/10.1016/j.jaad.2019.06.498**
**Three Phase 3 trials : B7451012, B7451013 and B7451029**
**https://www.accessdata.fda.gov/drugsatfda_docs/nda/2022/213871Orig1s000MultidisciplineR.pdf {page 89)**

| Drug | |
|---|---|
| Tradename | **Kimmtrak** |
| Generic Name | **TEBENTAFUSP-TEBN** |
| Company | Immunocore |
| Date of FDA Approval | January 25, 2022 |
| Indication | treatment of HLA-A*02:01-positive adult patients with unresectable or metastatic **uveal melanoma** |

**OVERALL GRADE**

**B**

### CARER Clinical Trial Diversity Scorecard

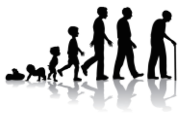

**Age**

| Age | % in U.S. Population | % in Clinical Trials | Number treated with new drug | Incidence of Disease or Condition | Grade |
|---|---|---|---|---|---|
| >/=65 years old | 16.5% | 49.5% | 122 | Increased | **B** |

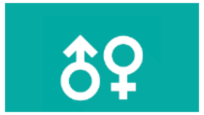

**Sex**

| Sex | % in U.S. Population | % in Clinical Trials | Number treated with new drug | Incidence of Disease or Condition | Grade |
|---|---|---|---|---|---|
| Female | 50.8% | 49.7% | 124 | Decreased | **A** |

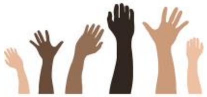

**Race**

| Race | % in U.S. Population | % in Clinical Trials | Number treated with new drug | Incidence of disease or condition | Grade |
|---|---|---|---|---|---|
| Black or African | 13.4% | 0% | 0 | Decreased | **C** |
| Asian | 5.9% | 0% | 0 | Decreased | **C** |

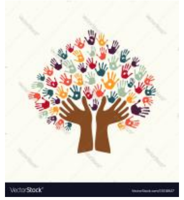

**Ethnicity**

| Ethnicity | % in U.S. Population | % in Clinical Trials | Number treated with new drug | Incidence of disease or condition | Grade |
|---|---|---|---|---|---|
| Hispanic or Latino | 18.5% | 2.4% | 3 | Decreased | **C** |


•**References:** doi: 10.1016/S0161-6420(03)00078-2; doi: 10.1136/bjo.2008.150292; DOI: 10.1016/j.ophtha.2011.01.040; doi: 10.1038/eye.2015.51.
**Phase 3 trial** IMCgp100-202
**https://www.accessdata.fda.gov/drugsatfda_docs/nda/2022/761228Orig1s000MultidisciplineR.pdf {page 99}**

| Drug | |
|---|---|
| Tradename | **Vabysmo** |
| Generic Name | **faricimab-svoa** |
| Company | Genentech |
| Date of FDA Approval | January 28, 2022 |
| Indication | To treat **diabetic macular edema** |

#### OVERALL GRADE

# B

### CARER Clinical Trial Diversity Scorecard

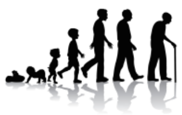

Age

| Age | % in U.S. Population | % in Clinical Trials | Number treated with new drug | Incidence of Disease or Condition | Grade |
|---|---|---|---|---|---|
| >/=65 years old | 16.5% | 42.9% | 548 | Similar | **A** |

Sex

| Sex | % in U.S. Population | % in Clinical Trials | Number treated with new drug | Incidence of Disease or Condition | Grade |
|---|---|---|---|---|---|
| Female | 50.8% | 39.7% | 487 | Similar | **A** |

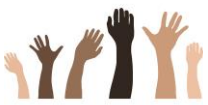

Race

| Race | % in U.S. Population | % in Clinical Trials | Number treated with new drug | Incidence of disease or condition | Grade |
|---|---|---|---|---|---|
| Black or African | 13.4% | 6.6% | 88 | Increased | **D** |
| Asian | 5.9% | 9.8% | 127 | Similar | **A** |

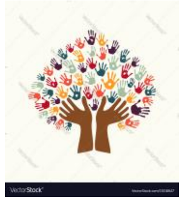

Ethnicity

| Ethnicity | % in U.S. Population | % in Clinical Trials | Number treated with new drug | Incidence of disease or condition | Grade |
|---|---|---|---|---|---|
| Hispanic or Latino | 18.5% | NR | NR | Similar | **D** |

**References: Phase 3 trials Yosemite, , ; https://www.accessdata.fda.gov/drugsatfda_docs/nda/2022/215833Orig1s000MultidisciplineR.pdf (page 67, 77); doi: 10.1001/jamaophthalmol.2014.2854**

| Drug | |
|---|---|
| Tradename | **Vabysmo** |
| Generic Name | **faricimab-svoa** |
| Company | Genentech |
| Date of FDA Approval | January 28, 2022 |
| Indication | To treat **Neovascular (Wet) Age-Related Macular Degeneration** |

#### OVERALL GRADE

# B

### CARER Clinical Trial Diversity Scorecard

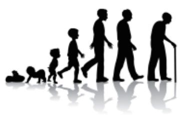

Age

| Age | % in U.S. Population | % in Clinical Trials | Number treated with new drug | Incidence of Disease or Condition | Grade |
|---|---|---|---|---|---|
| >/=75 years old | 7% | 59.3% | 379 | Increased | **A** |

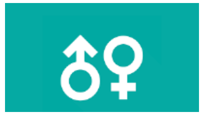

Sex

| Sex | % in U.S. Population | % in Clinical Trials | Number treated with new drug | Incidence of Disease or Condition | Grade |
|---|---|---|---|---|---|
| Female | 50.8% | 59.7% | 294 | Similar | **B** |

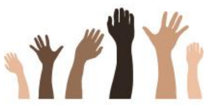

Race

| Race | % in U.S. Population | % in Clinical Trials | Number treated with new drug | Incidence of disease or condition | Grade |
|---|---|---|---|---|---|
| Black or African | 13.4% | 0.8% | 2 | Decreased | **C** |
| Asian | 5.9% | 9.5% | 64 | Decreased | **A** |

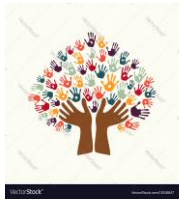

Ethnicity

| Ethnicity | % in U.S. Population | % in Clinical Trials | Number treated with new drug | Incidence of disease or condition | Grade |
|---|---|---|---|---|---|
| Hispanic or Latino | 18.5% | NR | NR | Similar | **D** |

**References: Phase 3 trials GR40306 (TENAYA), GR40844 (LUCERNE); https://www.accessdata.fda.gov/drugsatfda_docs/nda/2022/215833Orig1s000MultidisciplineR.pdf (page 31, 39); doi: 10.1186/s40662-016-0063-5;**

### CARER Clinical Trial Diversity Scorecard

| Drug | |
|---|---|
| Tradename | **Enjaymo** |
| Generic Name | **sutimlimab-jome** |
| Company | Bioverativ USA, Sanofi |
| Date of FDA Approval | February 4, 2022 |
| Indication | to decrease the need for red blood cell (RBC) transfusion due to hemolysis in adults with **cold agglutinin disease** (CAD**)** |

**OVERALL GRADE**

**C**

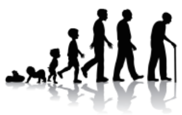

Age

| **Age** | **% in U.S. Population** | **% in Clinical Trials** | **Number treated with new drug** | **Incidence of Disease or Condition** | **Grade** |
|---|---|---|---|---|---|
| >/=65 years old | 16.5% | 79.2% | 19 | Increased | **C** |

Sex

| **Sex** | **% in U.S. Population** | **% in Clinical Trials** | **Number treated with new drug** | **Incidence of Disease or Condition** | **Grade** |
|---|---|---|---|---|---|
| Female | 50.8% | 62.5% | 15 | Increased | **D** |

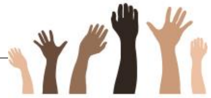

Race

| **Race** | **% in U.S. Population** | **% in Clinical Trials** | **Number treated with new drug** | **Incidence of disease or condition** | **Grade** |
|---|---|---|---|---|---|
| Black or African | 13.4% | 0% | 0 | Decreased | **C** |
| Asian | 5.9% | 12.5% | 3 | Similar | **B** |

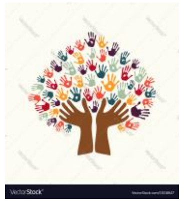

Ethnicity

| **Ethnicity** | **% in U.S. Population** | **% in Clinical Trials** | **Number treated with new drug** | **Incidence of disease or condition** | **Grade** |
|---|---|---|---|---|---|
| Hispanic or Latino | 18.5% | 0 | 0 | Decreased | **C** |

**References: https://doi.org/10.1182/blood.2020005674; https://doi.org/10.1182/blood-2013-02-474437; https://doi.org/10.1182/blood.V130.Suppl_1.928.928; Phase 3 Study BIVV009-03 Part A (CARDINAL); https://www.accessdata.fda.gov/drugsatfda_docs/nda/2022/761164Orig1s000MedR.pdf {page 39)**

### CARER Clinical Trial Diversity Scorecard

| Drug | |
|---|---|
| Tradename | **Pyrukynd** |
| Generic Name | **mitapivat** |
| Company | Agios Pharmaceuticals |
| Date of FDA Approval | February 17, 2022 |
| Indication | treatment of **hemolytic anemia in adults with pyruvate kinase (PK) deficiency** |

#### OVERALL GRADE

# C

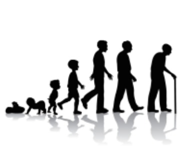

Age

| **Age** | **% in U.S. Population** | **% in Clinical Trials** | **Number treated with new drug** | **Incidence of Disease or Condition** | **Grade** |
|---|---|---|---|---|---|
| >/=65 years old | 16.5% | 4.7% | 3 | Decreased | **C** |

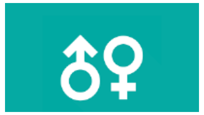

Sex

| **Sex** | **% in U.S. Population** | **% in Clinical Trials** | **Number treated with new drug** | **Incidence of Disease or Condition** | **Grade** |
|---|---|---|---|---|---|
| Female | 50.8% | 63.6% | 44 | Similar | **B** |

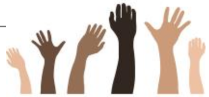

Race

| **Race** | **% in U.S. Population** | **% in Clinical Trials** | **Number treated with new drug** | **Incidence of disease or condition** | **Grade** |
|---|---|---|---|---|---|
| Black or African | 13.4% | 0% | 0 | Similar | **D** |
| Asian | 5.9% | 10.3% | 8 | Similar | **B** |

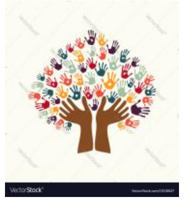

Ethnicity

| **Ethnicity** | **% in U.S. Population** | **% in Clinical Trials** | **Number treated with new drug** | **Incidence of disease or condition** | **Grade** |
|---|---|---|---|---|---|
| Hispanic or Latino | 18.5% | NR | NR | Decreased | **C** |

**References:** https://doi.org/10.1182/blood.V95.11.3585' n Study AG-348-C-006 (Study 006) Study AG-348-C-007 (Study 007); https://www.accessdata.fda.gov/drugsatfda_docs/nda/2022/216196Orig1s000IntegratedR.pdf **{page 35, 51}**

### CARER Clinical Trial Diversity Scorecard

| Drug | |
|---|---|
| Tradename | **Vonjo** |
| Generic Name | **pacritinib** |
| Company | CTI Biopharma |
| Date of FDA Approval | February 28, 2022 |
| Indication | treatment of adults with intermediate or high-risk primary or secondary (post-polycythemia vera or post-essential thrombocythemia) **myelofibrosis** with a platelet count below 50,000. This indication is approved under accelerated approval based on spleen volume reduction |

#### OVERALL GRADE

# C

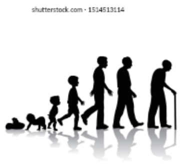


Age

| Age | % in U.S. Population | % in Clinical Trials | Number treated with new drug | Incidence of Disease or Condition | Grade |
|---|---|---|---|---|---|
| >/=65 years old | 16.5% | 62% | 99 | Increased | **B** |

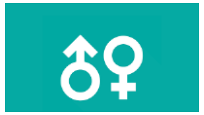

Sex

| Sex | % in U.S. Population | % in Clinical Trials | Number treated with new drug | Incidence of Disease or Condition | Grade |
|---|---|---|---|---|---|
| Female | 50.8% | 43% | 63 | Decreased | **B** |

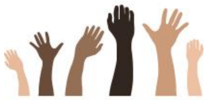

Race

| Race | % in U.S. Population | % in Clinical Trials | Number treated with new drug | Incidence of disease or condition | Grade |
|---|---|---|---|---|---|
| Black or African | 13.4% | 0.5% | 1 | Similar | **D** |
| Asian | 5.9% | 2% | 3 | Decreased | **C** |

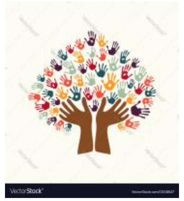

Ethnicity

| Ethnicity | % in U.S. Population | % in Clinical Trials | Number treated with new drug | Incidence of disease or condition | Grade |
|---|---|---|---|---|---|
| Hispanic or Latino | 18.5% | 3% | 3 | Decreased | **C** |

**References: doi:10.3324/haematol.2016.149559. PMC 5210236**; https://doi.org/10.1007/s00277-020-04055-w; https://doi.org/10.1016/j.hemonc.2021.01.005; doi: 10.1111/bjh.14061
**Phase 3 trial PERSIST-2**
**https://www.accessdata.fda.gov/drugsatfda_docs/nda/2022/208712Orig1s000IntegratedR.pdf {page 38}**

### CARER Clinical Trial Diversity Scorecard

| Drug | |
|---|---|
| Tradename | **Ztalmy** |
| Generic Name | **ganaxolone** |
| Company | Marinus |
| Date of FDA Approval | March 18, 2022 |
| Indication | treat **seizures in cyclin-dependent kinase-like 5 deficiency disorder in patients 2 years of age and older** |

#### OVERALL GRADE

# C

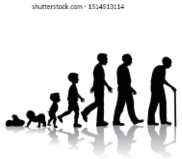

Age

| Age | % in U.S. Population | % in Clinical Trials | Number treated with new drug | Incidence of Disease or Condition | Grade |
|---|---|---|---|---|---|
| >/=65 years old | 16.5% | NA | NA | Decreased | **NA** |

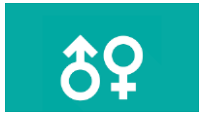

Sex

| Sex | % in U.S. Population | % in Clinical Trials | Number treated with new drug | Incidence of Disease or Condition | Grade |
|---|---|---|---|---|---|
| Female | 50.8% | 79.2% | 39 | Increased | **B** |

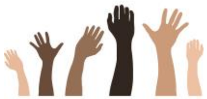

Race

| Race | % in U.S. Population | % in Clinical Trials | Number treated with new drug | Incidence of disease or condition | Grade |
|---|---|---|---|---|---|
| Black or African | 13.4% | NR | NR | Decreased | **C** |
| Asian | 5.9% | 5.0% | 2 | Similar | **C** |

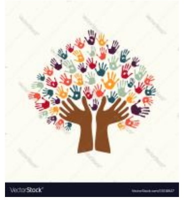

Ethnicity

| Ethnicity | % in U.S. Population | % in Clinical Trials | Number treated with new drug | Incidence of disease or condition | Grade |
|---|---|---|---|---|---|
| Hispanic or Latino | 18.5% | 9.9% | 4 | Similar | **D** |

**References: Phase 3 trial: 1042-CDD-3001; https://www.accessdata.fda.gov/drugsatfda_docs/nda/2022/215904Orig1s000MedR.pdf(page 42-43)); https://rarediseases.org/rare-diseases/cdkl5/; https://doi.org/10.1186/s11689-021-09384-z;**

| Drug | |
|---|---|
| Tradename | **Opdualag** |
| Generic Name | **nivolumab and relatlimab-rmbw** |
| Company | Bristol Myers Squibb |
| Date of FDA Approval | March 18, 2022 |
| Indication | To treat **unresectable or metastatic melanoma** |

**OVERALL GRADE**

**B**

### CARER Clinical Trial Diversity Scorecard

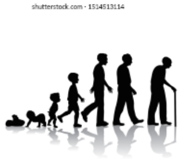

**Age**

| Age | % in U.S. Population | % in Clinical Trials | Number treated with new drug | Incidence of Disease or Condition | Grade |
|---|---|---|---|---|---|
| >/=65 years old | 16.5% | 46.4% | 168 | Increased | **B** |

**Sex**

| Sex | % in U.S. Population | % in Clinical Trials | Number treated with new drug | Incidence of Disease or Condition | Grade |
|---|---|---|---|---|---|
| Female | 50.8% | 41.7% | 145 | Decreased | **A** |

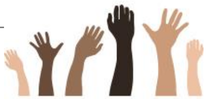

**Race**

| Race | % in U.S. Population | % in Clinical Trials | Number treated with new drug | Incidence of disease or condition | Grade |
|---|---|---|---|---|---|
| Black or African | 13.4% | 0.7% | 0 | Decreased | **C** |
| Asian | 5.9% | NR | NR | Decreased | **C** |

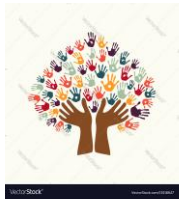

**Ethnicity**

| Ethnicity | % in U.S. Population | % in Clinical Trials | Number treated with new drug | Incidence of disease or condition | Grade |
|---|---|---|---|---|---|
| Hispanic or Latino | 18.5% | 6.6% | 27 | Decreased | **C** |

**References: https://www.cancer.org/cancer/melanoma-skin-cancer/about/key-statistics.html DOI:https://doi.org/10.1016/j.jaad.2020.08.086; Phase 3 Study CA224047 (RELATIVITY -047); https://www.accessdata.fda.gov/drugsatfda_docs/nda/2022/761234Orig1s000MultidisciplineR.pdf {page 132)**

| Drug | |
|---|---|
| Tradename | **Pluvitco** |
| Generic Name | **lutetium (177Lu) vipivotide tetraxetan** |
| Company | Advanced Accelerator Applications USA, Inc. |
| Date of FDA Approval | March 23, 2022 |
| Indication | To treat **prostate-specific membrane antigen-positive metastatic castration-resistant prostate cancer** following other therapies |

#### OVERALL GRADE

# C

### CARER Clinical Trial Diversity Scorecard

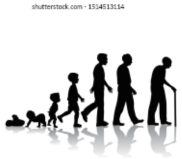

Age

| Age | % in U.S. Population | % in Clinical Trials | Number treated with new drug | Incidence of Disease or Condition | Grade |
|---|---|---|---|---|---|
| >/=65 years old | 16.5% | 75.3% | 406 | Increased | **A** |

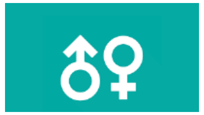

Sex

| Sex | % in U.S. Population | % in Clinical Trials | Number treated with new drug | Incidence of Disease or Condition | Grade |
|---|---|---|---|---|---|
| Female | 50.8% | NA | NA | Decreased | **NA** |

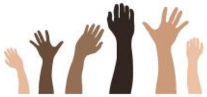

Race

| Race | % in U.S. Population | % in Clinical Trials | Number treated with new drug | Incidence of disease or condition | Grade |
|---|---|---|---|---|---|
| Black or African | 13.4% | 6.6% | 34 | Increased | **D** |
| Asian | 5.9% | 2.4% | 9 | Decreased | **C** |

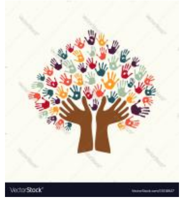

Ethnicity

| Ethnicity | % in U.S. Population | % in Clinical Trials | Number treated with new drug | Incidence of disease or condition | Grade |
|---|---|---|---|---|---|
| Hispanic or Latino | 18.5% | 1.7% | 11 | Decreased | **C** |

**References: Phase 3 trials PSMA-617-01; https://www.accessdata.fda.gov/drugsatfda_docs/nda/2022/215833Orig1s000MultidisciplineR.pdf (page 98 ); https://www.cancer.org/cancer/prostate-cancer/about/key-statistics.html#:~:text=Prostate%20cancer%20is%20more%20likely,at%20diagnosis%20is%20about%2066.; https://stacks.cdc.gov/view/cdc/94593**

| Drug | |
|---|---|
| Tradename | **Vivjoa** |
| Generic Name | **oteseconazole** |
| Company | Mycovia |
| Date of FDA Approval | April 26, 2022 |
| Indication | to reduce the incidence of **recurrent vulvovaginal candidiasis** (RVVC) in females with a history of RVVC who are NOT of reproductive potential |

### CARER Clinical Trial Diversity Scorecard

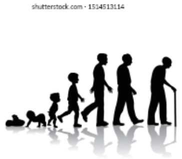

Age

| Age | % in U.S. Population | % in Clinical Trials | Number treated with new drug | Incidence of Disease or Condition | Grade |
|---|---|---|---|---|---|
| >/=65 years old | 16.5% | 0.6% | 3 | Decreased | **C** |

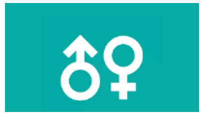

Sex

| Sex | % in U.S. Population | % in Clinical Trials | Number treated with new drug | Incidence of Disease or Condition | Grade |
|---|---|---|---|---|---|
| Female | 50.8% | 100% | 580 | Increased | **NA** |

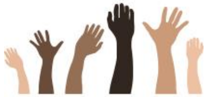

Race

| Race | % in U.S. Population | % in Clinical Trials | Number treated with new drug | Incidence of disease or condition | Grade |
|---|---|---|---|---|---|
| Black or African | 13.4% | 17.1% | 96 | Similar | **B** |
| Asian | 5.9% | 5.9% | 36 | Similar | **B** |

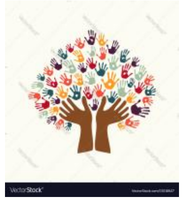

Ethnicity

| Ethnicity | % in U.S. Population | % in Clinical Trials | Number treated with new drug | Incidence of disease or condition | Grade |
|---|---|---|---|---|---|
| Hispanic or Latino | 18.5% | 14.8% | 86 | Similar | **B** |

#### OVERALL GRADE

# B

**References: Phase 3 trials: -CL-011, CL-012, CL-017;**
**https://www.accessdata.fda.gov/drugsatfda_docs/nda/2022/215888Orig1s000IntegratedR.pdf (page 255 ); doi: 10.1186/s12905-019-0748-8;**
**https://bmcwomenshealth.biomedcentral.com/articles/10.1186/s12905-022-01741-x/tables/4**

### CARER Clinical Trial Diversity Scorecard

| Drug | |
|---|---|
| Tradename | **Camzyos** |
| Generic Name | **mavacamten** |
| Company | Bristol Myers Squibb |
| Date of FDA Approval | April 28, 2022 |
| Indication | To treat certain classes of **obstructive hypertrophic cardiomyopathy** |

**OVERALL GRADE**

# D

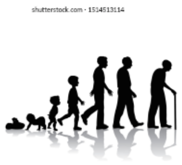

Age

| Age | % in U.S. Population | % in Clinical Trials | Number treated with new drug | Incidence of Disease or Condition | Grade |
|---|---|---|---|---|---|
| >/=65 years old | 16.5% | 33.9% | 45 | Increased | **B** |

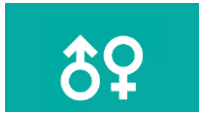

Sex

| Sex | % in U.S. Population | % in Clinical Trials | Number treated with new drug | Incidence of Disease or Condition | Grade |
|---|---|---|---|---|---|
| Female | 50.8% | 40.6% | 57 | Similar | **B** |

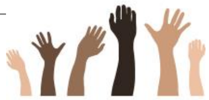

Race

| Race | % in U.S. Population | % in Clinical Trials | Number treated with new drug | Incidence of disease or condition | Grade |
|---|---|---|---|---|---|
| Black or African | 13.4% | 2.4% | 1 | Increased | **F** |
| Asian | 5.9% | 2.4% | 4 | Similar | **D** |

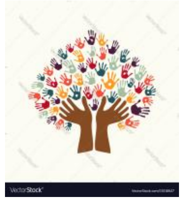

Ethnicity

| Ethnicity | % in U.S. Population | % in Clinical Trials | Number treated with new drug | Incidence of disease or condition | Grade |
|---|---|---|---|---|---|
| Hispanic or Latino | 18.5% | 4.8% | 8 | Similar | **D** |

**References: doi: 10.1155/2018/3750879; https://doi.org/10.1161/JAHA.119.014448; Phase 3 Study EXPLORER-HCM (NCT-03470545); hhttps://www.accessdata.fda.gov/drugsatfda_docs/nda/2022/214998Orig1s000Med_StatR.pdf (page 39)**

| Drug | |
|---|---|
| Tradename | **Voquezna** |
| Generic Name | **vonoprazan, amoxicillin, and clarithromycin** |
| Company | Phathom Pharmaceuticals |
| Date of FDA Approval | May 3, 2022 |
| Indication | To treat **Helicobacter pylori infection** |

### CARER Clinical Trial Diversity Scorecard

#### OVERALL GRADE

# B

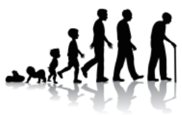

Age

| Age | % in U.S. Population | % in Clinical Trials | Number treated with new drug | Incidence of Disease or Condition | Grade |
|---|---|---|---|---|---|
| >/=65 years old | 16.5% | 19.9% | 153 | Increased | **C** |

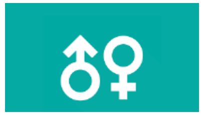

Sex

| Sex | % in U.S. Population | % in Clinical Trials | Number treated with new drug | Incidence of Disease or Condition | Grade |
|---|---|---|---|---|---|
| Female | 50.8% | 62.3% | 262 | Similar | **B** |

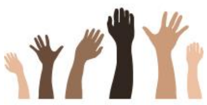

Race

| Race | % in U.S. Population | % in Clinical Trials | Number treated with new drug | Incidence of disease or condition | Grade |
|---|---|---|---|---|---|
| Black or African | 13.4% | 7.4% | 52 | Increased | **D** |
| Asian | 5.9% | 1.5% (100%)* | 10 (329)* | Similar | **A** |

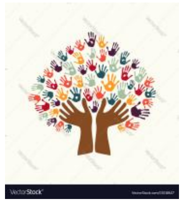

Ethnicity

| Ethnicity | % in U.S. Population | % in Clinical Trials | Number treated with new drug | Incidence of disease or condition | Grade |
|---|---|---|---|---|---|
| Hispanic or Latino | 18.5% | 27.1% | 194 | Increased | **B** |

**References: ; Phase 3 Study HP-301;*CCT-401 (Japan; all Asian); doi: 10.1086/315384 DOI: 10.1111/hel.12199;**
**https://wwwnc.cdc.gov/eid/article/10/6/03-0744_article**
**https://www.accessdata.fda.gov/drugsatfda_docs/nda/2022/215152Orig1s000,215153Orig1s000IntegratedR.pdf (page 25)**

| Drug | |
|---|---|
| Tradename | **Mounjaro** |
| Generic Name | **tirzepatide** |
| Company | Eli Lilly |
| Date of FDA Approval | May 13, 2022 |
| Indication | To improve blood sugar control in **diabetes**, in addition to diet and exercise |

### CARER Clinical Trial Diversity Scorecard

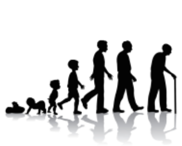

Age

| Age | % in U.S. Population | % in Clinical Trials | Number treated with new drug | Incidence of Disease or Condition | Grade |
|---|---|---|---|---|---|
| >/=65 years old | 16.5% | 31.8% | 1,466 | Increased | **A** |

Sex

| Sex | % in U.S. Population | % in Clinical Trials | Number treated with new drug | Incidence of Disease or Condition | Grade |
|---|---|---|---|---|---|
| Female | 50.8% | 43.3% | 2,166 | Similar | **A** |

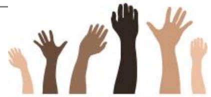

Race

| Race | % in U.S. Population | % in Clinical Trials | Number treated with new drug | Incidence of disease or condition | Grade |
|---|---|---|---|---|---|
| Black or African | 13.4% | 3.6% | 186 | Increased | **D** |
| Asian | 5.9% | 14.8% | 792 | Similar | **A** |

#### OVERALL GRADE

# B

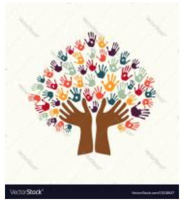

Ethnicity

| Ethnicity | % in U.S. Population | % in Clinical Trials | Number treated with new drug | Incidence of disease or condition | Grade |
|---|---|---|---|---|---|
| Hispanic or Latino | 18.5% | 42.3% | 2,041 | Increased | **A** |

**References: ; seven phase 3 trial (AS2); doi: 10.2147/IJGM.S226010; https://www.cdc.gov/diabetes/pdfs/library/diabetesreportcard2017-508.pdf https://www.accessdata.fda.gov/drugsatfda_docs/nda/2022/215866Orig1s000StatR.pdf(page 22)**

### CARER Clinical Trial Diversity Scorecard

| Drug | |
|---|---|
| Tradename | **Vtama** |
| Generic Name | **tapinarof** |
| Company | Dermavant |
| Date of FDA Approval | May 23, 2022 |
| Indication | To treat **plaque psoriasis** |

**OVERALL GRADE**

**A**

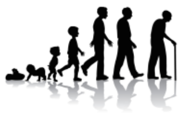

Age

| **Age** | **% in U.S. Population** | **% in Clinical Trials** | **Number treated with new drug** | **Incidence of Disease or Condition** | **Grade** |
|---|---|---|---|---|---|
| >/=65 years old | 16.5% | 13.6% | 99 | Similar | **B** |

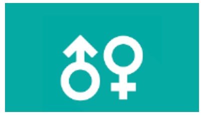

Sex

| **Sex** | **% in U.S. Population** | **% in Clinical Trials** | **Number treated with new drug** | **Incidence of Disease or Condition** | **Grade** |
|---|---|---|---|---|---|
| Female | 50.8% | 42.5% | 282 | Similar | **B** |

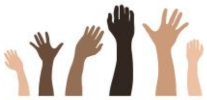

Race

| **Race** | **% in U.S. Population** | **% in Clinical Trials** | **Number treated with new drug** | **Incidence of disease or condition** | **Grade** |
|---|---|---|---|---|---|
| Black or African | 13.4% | 4.6% | 30 | Decreased | **B** |
| Asian | 5.9% | 6.9% | 46 | Decreased | **A** |

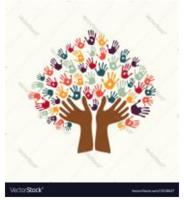

Ethnicity

| **Ethnicity** | **% in U.S. Population** | **% in Clinical Trials** | **Number treated with new drug** | **Incidence of disease or condition** | **Grade** |
|---|---|---|---|---|---|
| Hispanic or Latino | 18.5% | 14.9% | 96 | Decreased | **A** |

**References: Phase 3 trials: 3001, 3002; https://www.accessdata.fda.gov/drugsatfda_docs/nda/2022/215272Orig1s000MultidisciplineR.pdf(page 88-89); https://doi.org/10.1038/JID.2015.378; doi:10.1001/jamadermatol.2021.2007; Alexis AF, Blackcloud P. Psoriasis in skin of color: epidemiology, genetics, clinical presentation, and treatment nuances. J Clin Aesthet Dermatol. 2014 Nov;7(11):16-24. PMID: 25489378; PMCID: PMC4255694.**

| Drug | |
|---|---|
| Tradename | **Amvuttra** |
| Generic Name | **vutrisiran** |
| Company | Alnylam |
| Date of FDA Approval | June 13, 2022 |
| Indication | treatment of the **polyneuropathy of hereditary transthyretin-mediated amyloidosis** in adults |

#### OVERALL GRADE

# C

### CARER Clinical Trial Diversity Scorecard

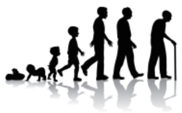

Age

| **Age** | **% in U.S. Population** | **% in Clinical Trials** | **Number treated with new drug** | **Incidence of Disease or Condition** | **Grade** |
|---|---|---|---|---|---|
| >/=65 years old | 16.5% | 34.8% | 46 | Similar | **A** |

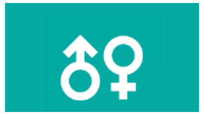

Sex

| **Sex** | **% in U.S. Population** | **% in Clinical Trials** | **Number treated with new drug** | **Incidence of Disease or Condition** | **Grade** |
|---|---|---|---|---|---|
| Female | 50.8% | 35.4% | 43 | Decreased | **B** |

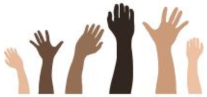

Race

| **Race** | **% in U.S. Population** | **% in Clinical Trials** | **Number treated with new drug** | **Incidence of disease or condition** | **Grade** |
|---|---|---|---|---|---|
| Black or African | 13.4% | 4.9% | 4 | Similar | **D** |
| Asian | 5.9% | 17.7% | 21 | Increased | **C** |

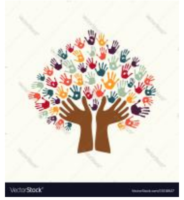

Ethnicity

| **Ethnicity** | **% in U.S. Population** | **% in Clinical Trials** | **Number treated with new drug** | **Incidence of disease or condition** | **Grade** |
|---|---|---|---|---|---|
| Hispanic or Latino | 18.5% | 9.8% | 12 | Decreased | **C** |

**References: Phase 3 trials: HELIOS-A (ALN-TTRSC02-002); https://www.accessdata.fda.gov/drugsatfda_docs/nda/2022/215515Orig1s000MedR.pdf (page 54-55); https://doi.org/10.1038/JID.2015.378; DOI: 10.1590/0004-282X-ANP-2020-0590; DOI https://doi.org/10.2147/TCRM.S219979**

| Drug | |
|---|---|
| Tradename | **Xenpozyme** |
| Generic Name | **Olipudase alfa** |
| Company | Genzyme |
| Date of FDA Approval | August 31, 2022 |
| Indication | To treat **Acid Sphingomyelinase Deficiency** |

#### OVERALL GRADE

# C

### CARER Clinical Trial Diversity Scorecard

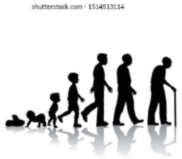

Age

| Age | % in U.S. Population | % in Clinical Trials | Number treated with new drug | Incidence of Disease or Condition | Grade |
|---|---|---|---|---|---|
| >/=65 years old | 16.5% | 1.8% | 0 | Decreased | **C** |

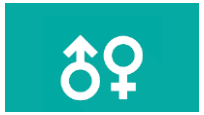

Sex

| Sex | % in U.S. Population | % in Clinical Trials | Number treated with new drug | Incidence of Disease or Condition | Grade |
|---|---|---|---|---|---|
| Female | 50.8% | 57.1% | 19 | Similar | **C** |

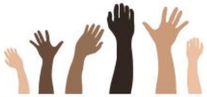

Race

| Race | % in U.S. Population | % in Clinical Trials | Number treated with new drug | Incidence of disease or condition | Grade |
|---|---|---|---|---|---|
| Black or African | 13.4% | 0% | 0 | Decreased | **C** |
| Asian | 5.9% | 7.1% | 3 | Similar | **C** |

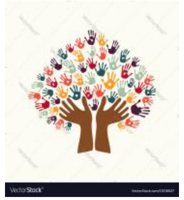

Ethnicity

| Ethnicity | % in U.S. Population | % in Clinical Trials | Number treated with new drug | Incidence of disease or condition | Grade |
|---|---|---|---|---|---|
| Hispanic or Latino | 18.5% | 21.4% | 6 | Increased | **C** |

**References: Phase 3 trials DFI12712 (ASCEND) DFI13803 (ASCEND-Peds);**
**https://www.accessdata.fda.gov/drugsatfda_docs/nda/2022/761261Orig1s000IntegratedR.pdf pg 38-39, 61-62, 92**
**https://doi.org/10.1016/j.jpeds.2006.06.034; https://rarediseases.org/rare-diseases/acid-sphingomyelinase-deficiency/;**
**https://www.ncbi.nlm.nih.gov/pmc/articles/PMC6122055/pdf/978-3-662-58081-3_Chapter_120.pdf;**

### CARER Clinical Trial Diversity Scorecard

| Drug | |
|---|---|
| Tradename | **Spevigo** |
| Generic Name | **Spesolimab-sbzo** |
| Company | Boehringer Ingelheim Pharmaceuticals |
| Date of FDA Approval | September 1, 2022 |
| Indication | To treat **generalized pustular psoriasis flares** |

#### OVERALL GRADE

# C

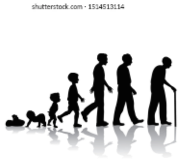

Age

| Age | % in U.S. Population | % in Clinical Trials | Number treated with new drug | Incidence of Disease or Condition | Grade |
|---|---|---|---|---|---|
| >/=65 years old | 16.5% | 4% | 2 | Similar | **D** |

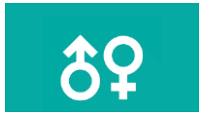

Sex

| Sex | % in U.S. Population | % in Clinical Trials | Number treated with new drug | Incidence of Disease or Condition | Grade |
|---|---|---|---|---|---|
| Female | 50.8% | 68% | 21 | Increased | **C** |

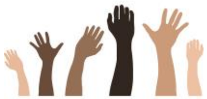

Race

| Race | % in U.S. Population | % in Clinical Trials | Number treated with new drug | Incidence of disease or condition | Grade |
|---|---|---|---|---|---|
| Black or African | 13.4% | 0% | 0 | Decreased | **C** |
| Asian | 5.9% | 55% | 16 | Increased | **C** |

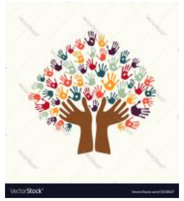

Ethnicity

| Ethnicity | % in U.S. Population | % in Clinical Trials | Number treated with new drug | Incidence of disease or condition | Grade |
|---|---|---|---|---|---|
| Hispanic or Latino | 18.5% | 0% | 0 | Decreased | **C** |

**References: Phase 3 trial Study Effisayil-1; https://www.accessdata.fda.gov/drugsatfda_docs/nda/2022/761244Orig1s000MultidisciplineR.pdf pg 72; https://doi.org/10.1080/1744666X.2019.1708193; doi:10.1001/jamadermatol.2021.2007; Alexis AF, Blackcloud P. Psoriasis in skin of color: epidemiology, genetics, clinical presentation, and treatment nuances. J Clin Aesthet Dermatol. 2014 Nov;7(11):16-24. PMID: 25489378; PMCID: PMC4255694.**

### CARER Clinical Trial Diversity Scorecard

| Drug | |
|---|---|
| Tradename | **Daxxify** |
| Generic Name | **daxibotulinumtoixnA-lanm** |
| Company | Revance Therapeutics |
| Date of FDA Approval | September 7, 2022 |
| Indication | To treat **moderate-to-severe glabellar lines** associated with corrugator and/or procerus muscle activity |

#### OVERALL GRADE

# B

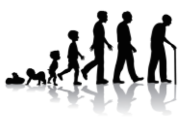

Age

| Age | % in U.S. Population | % in Clinical Trials | Number treated with new drug | Incidence of Disease or Condition | Grade |
|---|---|---|---|---|---|
| >/=55 years old | 29.1% | 30.4% | 105 | Increased | **C** |

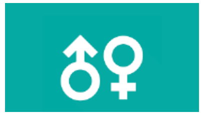

Sex

| Sex | % in U.S. Population | % in Clinical Trials | Number treated with new drug | Incidence of Disease or Condition | Grade |
|---|---|---|---|---|---|
| Female | 50.8% | 87.4% | 354 | Decreased | **A** |

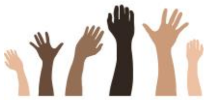

Race

| Race | % in U.S. Population | % in Clinical Trials | Number treated with new drug | Incidence of disease or condition | Grade |
|---|---|---|---|---|---|
| Black or African | 13.4% | 4.9% | 19 | Decreased | **C** |
| Asian | 5.9% | 2,6% | 18 | Decreased | **C** |

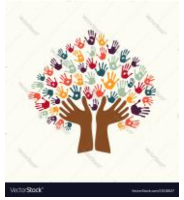

Ethnicity

| Ethnicity | % in U.S. Population | % in Clinical Trials | Number treated with new drug | Incidence of disease or condition | Grade |
|---|---|---|---|---|---|
| Hispanic or Latino | 18.5% | 16.6% | 66 | Decreased | **A** |

**References: Phase trials Study 301. 302 (GL1, GL2)**
**https://www.accessdata.fda.gov/drugsatfda_docs/nda/2022/761127Orig1s000MultidisciplineR.pdf pg 49 https://doi.org/10.1111/jocd.12806**
**https://jddonline.com/articles/ethnicity-and-aging-skin-S1545961617S0077X/; DOI: 10.1097/DSS.0000000000002237; DOI: 10.1111/dsu.12377**

### CARER Clinical Trial Diversity Scorecard

| Drug | |
|---|---|
| Tradename | **Rolvedon** |
| Generic Name | **eflapegrastim** |
| Company | Spectrum Pharmaceuticals |
| Date of FDA Approval | September 9, 2022 |
| Indication | To **decrease the incidence of infection** in patients with non-myeloid malignancies receiving myelosuppressive anti-cancer drugs associated with clinically significant incidence of febrile neutropenia |

#### OVERALL GRADE

# A

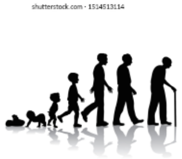

Age

| Age | % in U.S. Population | % in Clinical Trials | Number treated with new drug | Incidence of Disease or Condition | Grade |
|---|---|---|---|---|---|
| >/=65 years old | 16.5% | 37.8% | 122 | Increased | **B** |

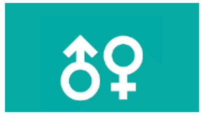

Sex

| Sex | % in U.S. Population | % in Clinical Trials | Number treated with new drug | Incidence of Disease or Condition | Grade |
|---|---|---|---|---|---|
| Female | 50.8% | 99.7% | 313 | Decreased | **A** |

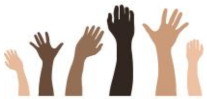

Race

| Race | % in U.S. Population | % in Clinical Trials | Number treated with new drug | Incidence of disease or condition | Grade |
|---|---|---|---|---|---|
| Black or African | 13.4% | 11.8% | 37 | Similar | **B** |
| Asian | 5.9% | 8.4% | 29 | Decreased | **A** |

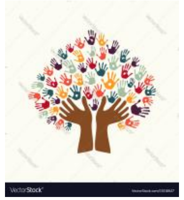

Ethnicity

| Ethnicity | % in U.S. Population | % in Clinical Trials | Number treated with new drug | Incidence of disease or condition | Grade |
|---|---|---|---|---|---|
| Hispanic or Latino | 18.5% | 16.6% | 52 | Decreased | **A** |

**References: Phase 3 trials SPI-GCF-301 and SPI-GCF-302 ; https://www.accessdata.fda.gov/drugsatfda_docs/nda/2022/761148Orig1s000MedR.pdf pp 44 ; https://seer.cancer.gov/statfacts/html/disparities.html**

| Drug | |
|---|---|
| Tradename | **Sotyktu** |
| Generic Name | **deucravacitinib** |
| Company | Bristol Myers Squibb |
| Date of FDA Approval | September 9, 2022 |
| Indication | To treat moderate-to-severe **plaque psoriasis** |

### CARER Clinical Trial Diversity Scorecard

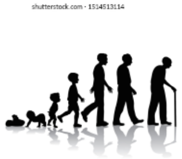

Age

| Age | % in U.S. Population | % in Clinical Trials | Number treated with new drug | Incidence of Disease or Condition | Grade |
|---|---|---|---|---|---|
| >/=65 years old | 16.5% | 10% | 80 | Similar | **C** |

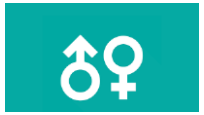

Sex

| Sex | % in U.S. Population | % in Clinical Trials | Number treated with new drug | Incidence of Disease or Condition | Grade |
|---|---|---|---|---|---|
| Female | 50.8% | 33% | 277 | Similar | **C** |

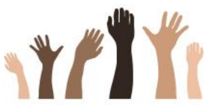

Race

| Race | % in U.S. Population | % in Clinical Trials | Number treated with new drug | Incidence of disease or condition | Grade |
|---|---|---|---|---|---|
| Black or African | 13.4% | 2% | 10 | Decreased | **C** |
| Asian | 5.9% | 10% | 83 | Decreased | **A** |

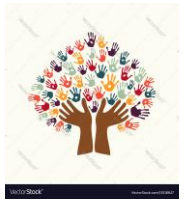

Ethnicity

| Ethnicity | % in U.S. Population | % in Clinical Trials | Number treated with new drug | Incidence of disease or condition | Grade |
|---|---|---|---|---|---|
| Hispanic or Latino | 18.5% | NR | NR | Decreased | **C** |

#### OVERALL GRADE

# C

**References: Phase 3 trials IM011046/47 (PSO-1, PSO-2); https://www.accessdata.fda.gov/drugsatfda_docs/nda/2022/214958Orig1s000MultidisciplineR.pdfhttps://doi.org/10.1038/JID.2015.378 (pg 121) ; doi:10.1001/jamadermatol.2021.2007; Alexis AF, Blackcloud P. Psoriasis in skin of color: epidemiology, genetics, clinical presentation, and treatment nuances. J Clin Aesthet Dermatol. 2014 Nov;7(11):16-24. PMID: 25489378; PMCID: PMC4255694.**

| Drug | |
|---|---|
| Tradename | **Terlivaz** |
| Generic Name | **terlipressin** |
| Company | Mallinckrodt |
| Date of FDA Approval | September 14, 2022 |
| Indication | To improve kidney function in adults with **hepatorenal syndrome** with rapid reduction in kidney function |

#### OVERALL GRADE

# C

### CARER Clinical Trial Diversity Scorecard

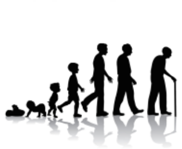

Age

| Age | % in U.S. Population | % in Clinical Trials | Number treated with new drug | Incidence of Disease or Condition | Grade |
|---|---|---|---|---|---|
| >/=65 years old | 16.5% | 17.7% | 35 | Increased | **C** |

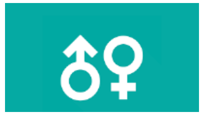

Sex

| Sex | % in U.S. Population | % in Clinical Trials | Number treated with new drug | Incidence of Disease or Condition | Grade |
|---|---|---|---|---|---|
| Female | 50.8% | 40.3% | 79 | Similar | **C** |

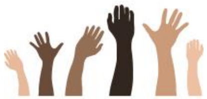

Race

| Race | % in U.S. Population | % in Clinical Trials | Number treated with new drug | Incidence of disease or condition | Grade |
|---|---|---|---|---|---|
| Black or African | 13.4% | 5.7% | 12 | Similar | **D** |
| Asian | 5.9% | 2.0% | 5 | Similar | **D** |

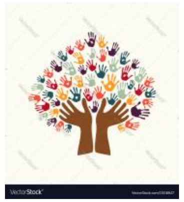

Ethnicity

| Ethnicity | % in U.S. Population | % in Clinical Trials | Number treated with new drug | Incidence of disease or condition | Grade |
|---|---|---|---|---|---|
| Hispanic or Latino | 18.5% | 15.0% | 32 | Similar | **B** |

**References: Phase 3 trial CONFIRM; https://www.accessdata.fda.gov/drugsatfda_docs/nda/2022/022231Orig1s000IntegratedR.pdf pg 26, 33 :https://rarediseases.org/rare-diseases/hepatorenal-syndrome/#:~:text=The%20exact%20incidence%20of%20hepatorenal,abdomen%20(ascites)%20and%20cirrhosis.10.1001/jamadermatol.2021.2007; https://www.wikidoc.org/index.php/Hepatorenal_syndrome_epidemiology_and_demographics**

### CARER Clinical Trial Diversity Scorecard

| Drug | |
|---|---|
| Tradename | **Elucirem** |
| Generic Name | **gadopiclenol** |
| Company | Guerbet |
| Date of FDA Approval | September 21 2022 |
| Indication | **To detect and visualize lesions, together with MRI, with abnormal vascularity in the central nervous system and the body** |

#### OVERALL GRADE

# B

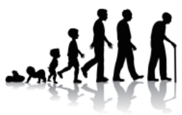

Age

| Age | % in U.S. Population | % in Clinical Trials | Number treated with new drug | Incidence of Disease or Condition | Grade |
|---|---|---|---|---|---|
| >/=65 years old | 16.5% | 34.3% | 189 | Increased | **B** |

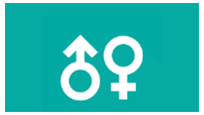

Sex

| Sex | % in U.S. Population | % in Clinical Trials | Number treated with new drug | Incidence of Disease or Condition | Grade |
|---|---|---|---|---|---|
| Female | 50.8% | 56.8% | 313 | Similar | **A** |

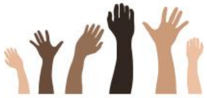

Race

| Race | % in U.S. Population | % in Clinical Trials | Number treated with new drug | Incidence of disease or condition | Grade |
|---|---|---|---|---|---|
| Black or African | 13.4% | 2.0% | 11 | Similar | **D** |
| Asian | 5.9% | 11.8% | 65 | Similar | **A** |

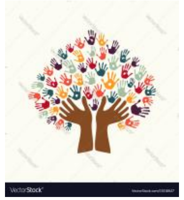

Ethnicity

| Ethnicity | % in U.S. Population | % in Clinical Trials | Number treated with new drug | Incidence of disease or condition | Grade |
|---|---|---|---|---|---|
| Hispanic or Latino | 18.5% | 12.5% | 69 | Similar | **C** |

**References: Phase 3 trials GDX-44-010 GDX-44-011**
https://www.accessdata.fda.gov/drugsatfda_docs/nda/2022/216986Orig1s000MultidisciplineR.pdf pg 117. **131**

### CARER Clinical Trial Diversity Scorecard

| Drug | |
|---|---|
| Tradename | **Omlonti** |
| Generic Name | **oomidenepag isopropyl ophthalmic solution** |
| Company | Santen |
| Date of FDA Approval | September 22, 2022 |
| Indication | To reduce elevated intraocular pressure in patients with **open-angle glaucoma or ocular hypertension** |

#### OVERALL GRADE

# B

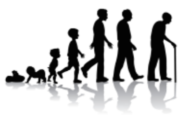

Age

| Age | % in U.S. Population | % in Clinical Trials | Number treated with new drug | Incidence of Disease or Condition | Grade |
|---|---|---|---|---|---|
| >/=65 years old | 16.5% | 46.6% | 285 | Increased | **B** |

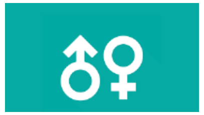

Sex

| Sex | % in U.S. Population | % in Clinical Trials | Number treated with new drug | Incidence of Disease or Condition | Grade |
|---|---|---|---|---|---|
| Female | 50.8% | 55.1% | 322 | Similar | **A** |

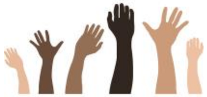

Race

| Race | % in U.S. Population | % in Clinical Trials | Number treated with new drug | Incidence of disease or condition | Grade |
|---|---|---|---|---|---|
| Black or African | 13.4% | 18.8% | 120 | Increased | **B** |
| Asian | 5.9% | 32.2% | 192 | Similar | **B** |

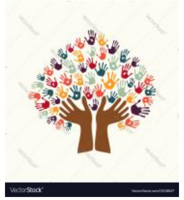

Ethnicity

| Ethnicity | % in U.S. Population | % in Clinical Trials | Number treated with new drug | Incidence of disease or condition | Grade |
|---|---|---|---|---|---|
| Hispanic or Latino | 18.5% | NR | NR | Increased | **F** |

**References: Phase 3 trials Studies 01171505, 011710IN, and 011709IN; https://www.accessdata.fda.gov/drugsatfda_docs/nda/2022/215092Orig1s000MedR.pdf pg 48; DOI: 10.1001/archopht.122.4.532 ; https://www.brightfocus.org/glaucoma/article/how-glaucoma-affects-different-ethnic-groups#:~:text=The%20prevalence%20of%20open%2Dangle,not%20know%20they%20had%20glaucoma.**

| Drug | |
|---|---|
| Tradename | **Relyvrio** |
| Generic Name | **sodium phenylbutyrate/taurursodiol** |
| Company | Amylyx Pharmaceuticals |
| Date of FDA Approval | September 29, 2022 |
| Indication | To treat **amyotrophic lateral sclerosis (ALS)** |

### CARER Clinical Trial Diversity Scorecard

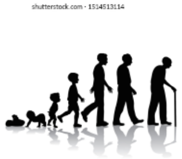

Age

| Age | % in U.S. Population | % in Clinical Trials | Number treated with new drug | Incidence of Disease or Condition | Grade |
|---|---|---|---|---|---|
| >/=65 years old | 16.5% | 23.4% | 25 | Increased | **C** |

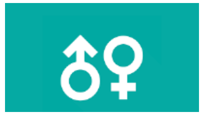

Sex

| Sex | % in U.S. Population | % in Clinical Trials | Number treated with new drug | Incidence of Disease or Condition | Grade |
|---|---|---|---|---|---|
| Female | 50.8% | 32.1% | 28 | Decreased | **C** |

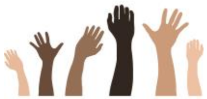

Race

| Race | % in U.S. Population | % in Clinical Trials | Number treated with new drug | Incidence of disease or condition | Grade |
|---|---|---|---|---|---|
| Black or African | 13.4% | 2.2% | 2 | Decreased | **C** |
| Asian | 5.9% | 2.2% | 2 | Decreased | **C** |

#### OVERALL GRADE

# C

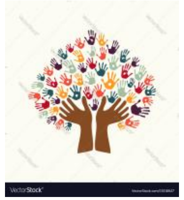

Ethnicity

| Ethnicity | % in U.S. Population | % in Clinical Trials | Number treated with new drug | Incidence of disease or condition | Grade |
|---|---|---|---|---|---|
| Hispanic or Latino | 18.5% | 5.1% | 6 | Decreased | **C** |

**References: Phase 3 trial Study 1; NCT03127514; https://www.accessdata.fda.gov/drugsatfda_docs/nda/2022/216660Orig1s000Med_StatR.pdf p 47; https://www.als.org/understanding-als/who-gets-als#:~:text=Most%20people%20who%20develop%20ALS,common%20in%20men%20than%20women. doi: 10.3109/21678421.2014.971813. https://www.cdc.gov/mmwr/volumes/67/wr/mm6707a3.htm**

### CARER Clinical Trial Diversity Scorecard

| Drug | |
|---|---|
| Tradename | **Lytgobi** |
| Generic Name | **futibatinib** |
| Company | Taiho Oncology |
| Date of FDA Approval | September 30, 2022 |
| Indication | To treat intrahepatic **cholangiocarcinoma** harboring fibroblast growth factor receptor 2 (FGFR2) gene fusions or other rearrangements |

#### OVERALL GRADE

# C

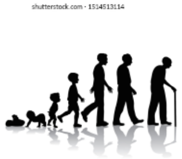
Age

| Age | % in U.S. Population | % in Clinical Trials | Number treated with new drug | Incidence of Disease or Condition | Grade |
|---|---|---|---|---|---|
| >/=65 years old | 16.5% | 22.3% | 23 | Increased | **C** |

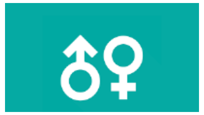
Sex

| Sex | % in U.S. Population | % in Clinical Trials | Number treated with new drug | Incidence of Disease or Condition | Grade |
|---|---|---|---|---|---|
| Female | 50.8% | 56.3 | 58 | Similar | **B** |

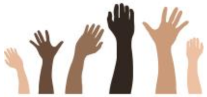
Race

| Race | % in U.S. Population | % in Clinical Trials | Number treated with new drug | Incidence of disease or condition | Grade |
|---|---|---|---|---|---|
| Black or African | 13.4% | 7.8% | 8 | Decreased | **C** |
| Asian | 5.9% | 29.1% | 30 | Increased | **B** |

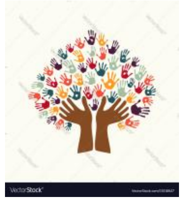
Ethnicity

| Ethnicity | % in U.S. Population | % in Clinical Trials | Number treated with new drug | Incidence of disease or condition | Grade |
|---|---|---|---|---|---|
| Hispanic or Latino | 18.5% | 1.9% | 2 | Increased | **F** |

**References: Phase 3 trial TAS-120-101; ; https://www.accessdata.fda.gov/drugsatfda_docs/nda/2022/214801Orig1s000MultidisciplineR.pdf pp 99 https://www.cancer.org/cancer/bile-duct-cancer/about/key-statistics.html; DOI:10.5604/01.3001.0010.8663; https://doi.org/10.1016/j.gastha.2021.12.003**

| Drug | |
|---|---|
| Tradename | **Imjudo** |
| Generic Name | **tremelimumab** |
| Company | AstraZeneca |
| Date of FDA Approval | October 21, 2022 |
| Indication | To treat unresectable **hepatocellular carcinoma** |

#### OVERALL GRADE

# C

### CARER Clinical Trial Diversity Scorecard

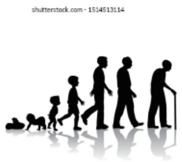


Age

| Age | % in U.S. Population | % in Clinical Trials | Number treated with new drug | Incidence of Disease or Condition | Grade |
|---|---|---|---|---|---|
| >/=65 years old | 16.5% | 50.6% | 277 | Increased | **B** |

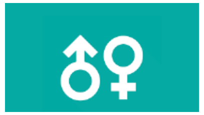

Sex

| Sex | % in U.S. Population | % in Clinical Trials | Number treated with new drug | Incidence of Disease or Condition | Grade |
|---|---|---|---|---|---|
| Female | 50.8% | 16.3% | 98 | Decreased | **B** |

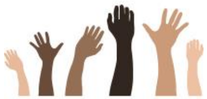

Race

| Race | % in U.S. Population | % in Clinical Trials | Number treated with new drug | Incidence of disease or condition | Grade |
|---|---|---|---|---|---|
| Black or African | 13.4% | 1.7% | 11 | Increased | **F** |
| Asian | 5.9% | 50.7% | 270 | Increased | **B** |

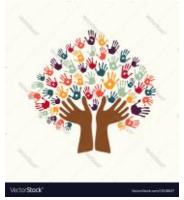

Ethnicity

| Ethnicity | % in U.S. Population | % in Clinical Trials | Number treated with new drug | Incidence of disease or condition | Grade |
|---|---|---|---|---|---|
| Hispanic or Latino | 18.5% | 5.0% | 32 | Increased | **D** |

**References: Phase 3 trial HIMALAYA; https://www.accessdata.fda.gov/drugsatfda_docs/nda/2022/761289Orig1s000MultidisciplineR.pdf pg 91 https://seer.cancer.gov/statfacts/html/livibd.html; https://doi.org/10.1002/hep4.1575**

| Drug | |
|---|---|
| Tradename | **Tecvayli** |
| Generic Name | **teclistamab-cqyv** |
| Company | Janssen |
| Date of FDA Approval | October 25, 2022 |
| Indication | To treat **relapsed or refractory multiple myeloma** among adults who have received at least four specific lines of therapy |

### CARER Clinical Trial Diversity Scorecard

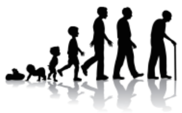

Age

| Age | % in U.S. Population | % in Clinical Trials | Number treated with new drug | Incidence of Disease or Condition | Grade |
|---|---|---|---|---|---|
| >/=65 years old | 16.5% | 48.8% | 78 | Increased | **B** |

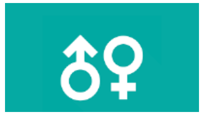

Sex

| Sex | % in U.S. Population | % in Clinical Trials | Number treated with new drug | Incidence of Disease or Condition | Grade |
|---|---|---|---|---|---|
| Female | 50.8% | 40.6% | 65 | Similar | **B** |

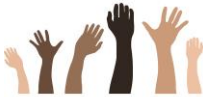

Race

| Race | % in U.S. Population | % in Clinical Trials | Number treated with new drug | Incidence of disease or condition | Grade |
|---|---|---|---|---|---|
| Black or African | 13.4% | 10.0% | 16 | Increased | **D** |
| Asian | 5.9% | 1.9% | 3 | Decreased | **C** |

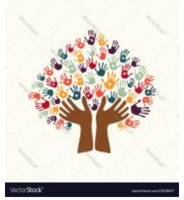

Ethnicity

| Ethnicity | % in U.S. Population | % in Clinical Trials | Number treated with new drug | Incidence of disease or condition | Grade |
|---|---|---|---|---|---|
| Hispanic or Latino | 18.5% | 9.4% | 15 | Similar | **D** |

#### OVERALL GRADE

# C

**References: Phase 3 trial -MajesTEC-1, NCT03145181 [Phase 1] and NCT04557098 [Phase 2]; (Study 64007957MMY1001; Pivotal RP2D); https://www.accessdata.fda.gov/drugsatfda_docs/nda/2022/761291Orig1s000MultidisciplineR.pdf pg 184 , RP2D SC (Part 1/2+Part 3A) (n=160) https://www.cancer.org/cancer/multiple-myeloma/causes-risks-prevention/risk-factors.html; https://seer.cancer.gov/statfacts/html/mulmy.html**

| Drug | |
|---|---|
| Tradename | **Elahere** |
| Generic Name | **minretumomab soravtansine-gynx** |
| Company | ImmunoGen |
| Date of FDA Approval | November 14, 2022 |
| Indication | To treat patients with **recurrent ovarian cancer** that is resistant to platinum therapy |

#### OVERALL GRADE

# C

### CARER Clinical Trial Diversity Scorecard

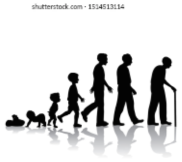

Age

| Age | % in U.S. Population | % in Clinical Trials | Number treated with new drug | Incidence of Disease or Condition | Grade |
|---|---|---|---|---|---|
| >/=65 years old | 16.5% | 43% | 45 | Increased | **B** |

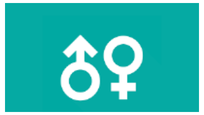

Sex

| Sex | % in U.S. Population | % in Clinical Trials | Number treated with new drug | Incidence of Disease or Condition | Grade |
|---|---|---|---|---|---|
| Female | 50.8% | 100% | 104 | Increased | **NA** |

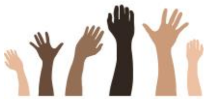

Race

| Race | % in U.S. Population | % in Clinical Trials | Number treated with new drug | Incidence of disease or condition | Grade |
|---|---|---|---|---|---|
| Black or African | 13.4% | 0% | 0 | Decreased | **C** |
| Asian | 5.9% | 2% | 2 | Decreased | **C** |

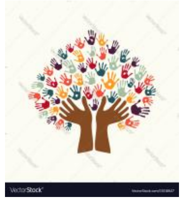

Ethnicity

| Ethnicity | % in U.S. Population | % in Clinical Trials | Number treated with new drug | Incidence of disease or condition | Grade |
|---|---|---|---|---|---|
| Hispanic or Latino | 18.5% | 2% | 2 | Similar | **D** |

**References: Phase 3 trial Study 0417; https://www.accessdata.fda.gov/drugsatfda_docs/nda/2022/761310Orig1s000MultidisciplineR.pdf pg 109 https://seer.cancer.gov/statfacts/html/ovary.html; https://doi.org/10.1002/cncr.11349; https://gis.cdc.gov/Cancer/USCS/#/Demographics/**

### CARER Clinical Trial Diversity Scorecard

| Drug | |
|---|---|
| Tradename | **Tzield** |
| Generic Name | **teplizumab-mzwv** |
| Company | Provention Bio |
| Date of FDA Approval | November 18, 2022 |
| Indication | To delay the onset of stage 3 **type 1 diabetes** |

**OVERALL GRADE**

**C**

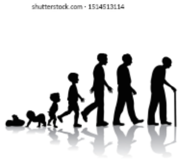

Age

| Age | % in U.S. Population | % in Clinical Trials | Number treated with new drug | Incidence of Disease or Condition | Grade |
|---|---|---|---|---|---|
| >/=65 years old | 16.5% | 0 | 0 | Decreased | **C** |

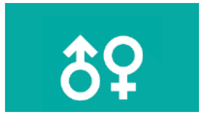

Sex

| Sex | % in U.S. Population | % in Clinical Trials | Number treated with new drug | Incidence of Disease or Condition | Grade |
|---|---|---|---|---|---|
| Female | 50.8% | 44.7% | 19 | Decreased | **B** |

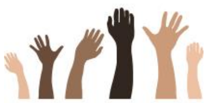

Race

| Race | % in U.S. Population | % in Clinical Trials | Number treated with new drug | Incidence of disease or condition | Grade |
|---|---|---|---|---|---|
| Black or African | 13.4% | 0 | 0 | Decreased | **C** |
| Asian | 5.9% | 1.3% | 0 | Decreased | **C** |

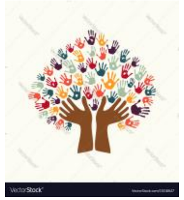

Ethnicity

| Ethnicity | % in U.S. Population | % in Clinical Trials | Number treated with new drug | Incidence of disease or condition | Grade |
|---|---|---|---|---|---|
| Hispanic or Latino | 18.5% | 2.6% | 2 | Decreased | **C** |

**References: Phase 3 trial Study TN-10l; https://www.accessdata.fda.gov/drugsatfda_docs/nda/2023/761183Orig1s000MedR.pdf pg 69; https://doi.org/10.1136/bmj.k1497**; https://doi.org/10.1186/s12916-017-0958-6; **doi:10.1056/NEJMoa1610187; doi: 10.1007/s001250051573; file:///C:/Users/billf/Downloads/NIHMS1052607-supplement-Supplementary_Appendix%20(1).pdf (non-Hispanic)**

| Drug | |
|---|---|
| Tradename | **Rezlidhia** |
| Generic Name | **olutasidenib** |
| Company | Rigel Pharmaceuticals |
| Date of FDA Approval | December 1, 2022 |
| Indication | To treat adults **with relapsed or refractory acute myeloid leukemia** with a susceptible isocitrate dehydrogenase-1 (IDH1) mutation |

#### OVERALL GRADE

# C

### CARER Clinical Trial Diversity Scorecard

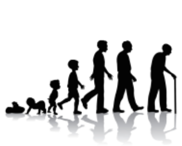

Age

| **Age** | **% in U.S. Population** | **% in Clinical Trials** | **Number treated with new drug** | **Incidence of Disease or Condition** | **Grade** |
|---|---|---|---|---|---|
| >/=65 years old | 16.5% | 75% | 116 | Increased | **B** |

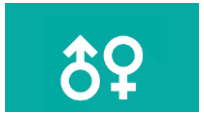

Sex

| **Sex** | **% in U.S. Population** | **% in Clinical Trials** | **Number treated with new drug** | **Incidence of Disease or Condition** | **Grade** |
|---|---|---|---|---|---|
| Female | 50.8% | 48% | 74 | Decreased | **A** |

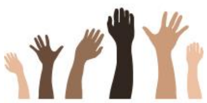

Race

| **Race** | **% in U.S. Population** | **% in Clinical Trials** | **Number treated with new drug** | **Incidence of disease or condition** | **Grade** |
|---|---|---|---|---|---|
| Black or African | 13.4% | 3% | 5 | Similar | **D** |
| Asian | 5.9% | 3% | 5 | Similar | **D** |

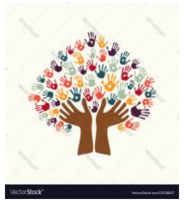

Ethnicity

| **Ethnicity** | **% in U.S. Population** | **% in Clinical Trials** | **Number treated with new drug** | **Incidence of disease or condition** | **Grade** |
|---|---|---|---|---|---|
| Hispanic or Latino | 18.5% | 5% | 8 | Similar | **D** |

**References: Phase 3 trial Study 2102-HEM-101; https://www.leukaemia.org.au/blood-cancer/leukaemia/acute-myeloid-leukaemia/; https://seer.cancer.gov/statfacts/html/amyl.html https://www.accessdata.fda.gov/drugsatfda_docs/nda/2022/215814Orig1s000MultidisciplineR.pdf pg 111;**

### CARER Clinical Trial Diversity Scorecard

| Drug | |
|---|---|
| Tradename | **Krazati** |
| Generic Name | **adagrasib** |
| Company | Mirati Therapeutics |
| Date of FDA Approval | December 12, 2022 |
| Indication | To treat KRAS G12C-mutated locally advanced or metastatic **non-small cell lung cancer** in adults who have received at least one prior systemic therapy |

#### OVERALL GRADE

# C

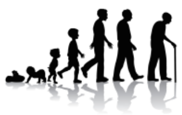

Age

| Age | % in U.S. Population | % in Clinical Trials | Number treated with new drug | Incidence of Disease or Condition | Grade |
|---|---|---|---|---|---|
| >/=65 years old | 16.5% | 47% | 53 | Increased | **B** |

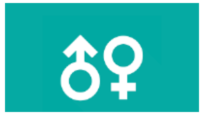

Sex

| Sex | % in U.S. Population | % in Clinical Trials | Number treated with new drug | Incidence of Disease or Condition | Grade |
|---|---|---|---|---|---|
| Female | 50.8% | 55% | 62 | Similar | **B** |

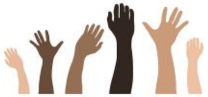

Race

| Race | % in U.S. Population | % in Clinical Trials | Number treated with new drug | Incidence of disease or condition | Grade |
|---|---|---|---|---|---|
| Black or African | 13.4% | 8% | 9 | Similar | **D** |
| Asian | 5.9% | 4.5% | 5 | Decreased | **B** |

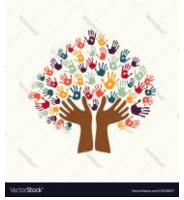

Ethnicity

| Ethnicity | % in U.S. Population | % in Clinical Trials | Number treated with new drug | Incidence of disease or condition | Grade |
|---|---|---|---|---|---|
| Hispanic or Latino | 18.5% | 2.7% | 3 | Decreased | **C** |

**References: Phase 3 trial KRYSTAL-1 ; https://www.accessdata.fda.gov/drugsatfda_docs/nda/2023/216340Orig1s000MultidisciplineR.pdf pg 120; DOI: 10.1056/NEJMc2030638 ; Cancer Control. 2016 October ; 23(4): 338–346; although females have lower incidence of NSLC, they have increased KRAS mutations in Whites=overall similar**

### CARER Clinical Trial Diversity Scorecard

| Drug | |
|---|---|
| Tradename | **Lunsumio** |
| Generic Name | **mosunetuzumab-axgb** |
| Company | Genentech |
| Date of FDA Approval | December 22, 2022 |
| Indication | To treat adults with **relapsed or refractory follicular lymphoma**, a type of non-Hodgkin lymphoma |

#### OVERALL GRADE

# B

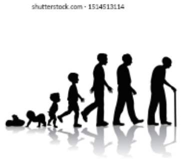

Age

| Age | % in U.S. Population | % in Clinical Trials | Number treated with new drug | Incidence of Disease or Condition | Grade |
|---|---|---|---|---|---|
| >/=65 years old | 16.5% | 33% | 30 | Increased | **B** |

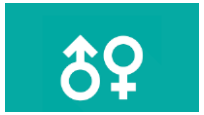

Sex

| Sex | % in U.S. Population | % in Clinical Trials | Number treated with new drug | Incidence of Disease or Condition | Grade |
|---|---|---|---|---|---|
| Female | 50.8% | 39% | 35 | Similar | **B** |

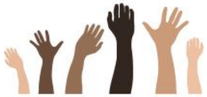

Race

| Race | % in U.S. Population | % in Clinical Trials | Number treated with new drug | Incidence of disease or condition | Grade |
|---|---|---|---|---|---|
| Black or African | 13.4% | 4% | 4 | Decreased | **C** |
| Asian | 5.9% | 9% | 8 | Decreased | **A** |

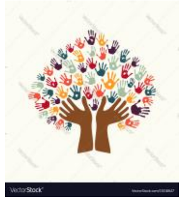

Ethnicity

| Ethnicity | % in U.S. Population | % in Clinical Trials | Number treated with new drug | Incidence of disease or condition | Grade |
|---|---|---|---|---|---|
| Hispanic or Latino | 18.5% | 8% | 7 | Similar | **D** |

**References: Phase 3 trial GO29781; https://seer.cancer.gov/statfacts/html/follicular.html; doi:10.1016/j.hoc.2020.02.001 ;**

### CARER Clinical Trial Diversity Scorecard

| Drug | |
|---|---|
| Tradename | **Sunlenca** |
| Generic Name | **lenacapavir** |
| Company | Gilead |
| Date of FDA Approval | December 22, 2022 |
| Indication | To treat adults with **HIV** whose HIV infections cannot be successfully treated with other available treatments due to resistance, intolerance, or safety considerations |

#### OVERALL GRADE

# B

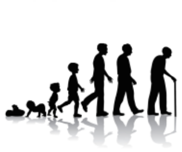

Age

| **Age** | **% in U.S. Population** | **% in Clinical Trials** | **Number treated with new drug** | **Incidence of Disease or Condition** | **Grade** |
|---|---|---|---|---|---|
| >/=50 years old | 34% | 75.0% | 16 | Decreased | **A** |

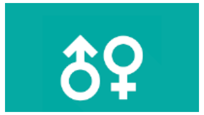

Sex

| **Sex** | **% in U.S. Population** | **% in Clinical Trials** | **Number treated with new drug** | **Incidence of Disease or Condition** | **Grade** |
|---|---|---|---|---|---|
| Female | 50.8% | 25.0% | 15 | Decreased | **C** |

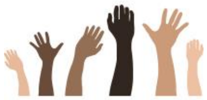

Race

| **Race** | **% in U.S. Population** | **% in Clinical Trials** | **Number treated with new drug** | **Incidence of disease or condition** | **Grade** |
|---|---|---|---|---|---|
| Black or African | 13.4% | 37.5% | 21 | Increased | **C** |
| Asian | 5.9% | 20.8% | 14 | Decreased | **A** |

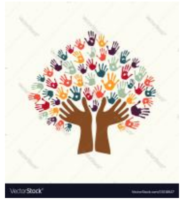

Ethnicity

| **Ethnicity** | **% in U.S. Population** | **% in Clinical Trials** | **Number treated with new drug** | **Incidence of disease or condition** | **Grade** |
|---|---|---|---|---|---|
| Hispanic or Latino | 18.5% | 21.5% | NR | Increased | **D** |

**References: Phase 3 trial CAPELLA 4625 ; https://www.gileadhiv.com/landscape/state-of-epidemic/?gclid=CjwKCAiAzKqdBhAnEiwAePEjkn-NINPZDHcdpXKfkXq9yiN1R8-XbjlFnUMnLXcLE-rY_WO966cnchoCC6EQAvD_BwE&gclsrc=aw.ds ; https://www.accessdata.fda.gov/drugsatfda_docs/label/2022/215973s000lbl.pdf; https://www.accessdata.fda.gov/drugsatfda_docs/nda/2023/215973,215974Orig1s000IntegratedR.pdf pg 29 https://www.jchs.harvard.edu/sites/default/files/jchs-housing_americas_older_adults_2014-ch2_0.pdf**

| Drug | |
|---|---|
| Tradename | **Xenoview** |
| Generic Name | **hyperpolarized Xe-129** |
| Company | Polarean |
| Date of FDA Approval | December 23, 2022 |
| Indication | To **evaluate pulmonary function and imaging** |

#### OVERALL GRADE

# C

### CARER Clinical Trial Diversity Scorecard

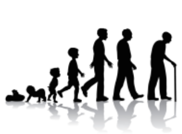

Age

| Age | % in U.S. Population | % in Clinical Trials | Number treated with new drug | Incidence of Disease or Condition | Grade |
|---|---|---|---|---|---|
| >/=65 years old | 16.5% | 44.4% | 36 | Increased | **B** |

Sex

| Sex | % in U.S. Population | % in Clinical Trials | Number treated with new drug | Incidence of Disease or Condition | Grade |
|---|---|---|---|---|---|
| Female | 50.8% | 30.9% | 25 | Similar | **D** |

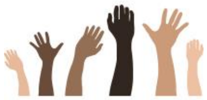

Race

| Race | % in U.S. Population | % in Clinical Trials | Number treated with new drug | Incidence of disease or condition | Grade |
|---|---|---|---|---|---|
| Black or African | 13.4% | 11.1% | 9 | Decreased | **B** |
| Asian | 5.9% | 1.2% | 1 | Decreased | **C** |

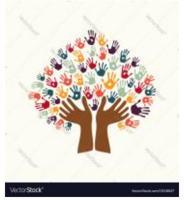

Ethnicity

| Ethnicity | % in U.S. Population | % in Clinical Trials | Number treated with new drug | Incidence of disease or condition | Grade |
|---|---|---|---|---|---|
| Hispanic or Latino | 18.5% | 1.2% | 1 | Decreased | **C** |

**References: Phase 3 trials Study POL-Xe-001; POL-Xe-002; https://www.accessdata.fda.gov/drugsatfda_docs/nda/2023/214375Orig1s000MultidisciplineR.pdf pg 50,58 ; Xenoview USPI for age >65yo; DOI: 10.1183/23120541.00630-2021 ; doi:10.1016/j.rmed.2012.01.002. https://www.lung.org/research/trends-in-lung-disease/copd-trends-brief/copd-prevalence; DOI https://doi.org/10.2147/COPD.S96391**

### CARER Clinical Trial Diversity Scorecard

| Drug | |
|---|---|
| Tradename | **NexoBrid** |
| Generic Name | **anacaulase-bcdb** |
| Company | MediWound |
| Date of FDA Approval | December 28, 2022 |
| Indication | **To remove eschar** in adults with deep partial thickness or full thickness **thermal burns** |

#### OVERALL GRADE

# D

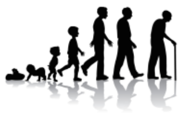

Age

| Age | % in U.S. Population | % in Clinical Trials | Number treated with new drug | Incidence of Disease or Condition | Grade |
|---|---|---|---|---|---|
| >/=65 years old | 16.5% | 4.8% | 6 | Decreased | **C** |

Sex

| Sex | % in U.S. Population | % in Clinical Trials | Number treated with new drug | Incidence of Disease or Condition | Grade |
|---|---|---|---|---|---|
| Female | 50.8% | 25.1% | 47 | Decreased | **B** |

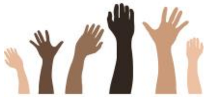

Race

| Race | % in U.S. Population | % in Clinical Trials | Number treated with new drug | Incidence of disease or condition | Grade |
|---|---|---|---|---|---|
| Black or African | 13.4% | 10.0% | 12 | Increased | **D** |
| Asian | 5.9% | 3.0% | 6 | Similar | **D** |

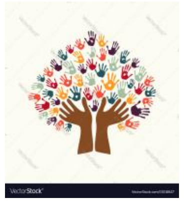

Ethnicity

| Ethnicity | % in U.S. Population | % in Clinical Trials | Number treated with new drug | Incidence of disease or condition | Grade |
|---|---|---|---|---|---|
| Hispanic or Latino | 18.5% | 9.1% | 14 | Similar | **D** |

**References: Phase 3 trials 2010, 2004; https://www.accessdata.fda.gov/drugsatfda_docs/nda/2023/761192Orig1s000MultidisciplineR.pdf pgs 83, 112; https://ameriburn.org/who-we-are/media/burn-incidence-fact-sheet/ ; https://www.researchgate.net/figure/Age-distribution-of-burned-patients_fig1_269185690**

### CARER Clinical Trial Diversity Scorecard

| Drug | |
|---|---|
| Tradename | **Briumvi** |
| Generic Name | **ublituximab-xiiy** |
| Company | TG Therapeutics |
| Date of FDA Approval | December 28, 2022 |
| Indication | To treat relapsing forms of **multiple sclerosis** |

#### OVERALL GRADE

# C

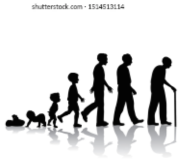

Age

| Age | % in U.S. Population | % in Clinical Trials | Number treated with new drug | Incidence of Disease or Condition | Grade |
|---|---|---|---|---|---|
| >/=40 years old | 49.3% | 35.0% | 172 | Decreased | **A** |

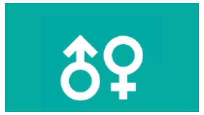

Sex

| Sex | % in U.S. Population | % in Clinical Trials | Number treated with new drug | Incidence of Disease or Condition | Grade |
|---|---|---|---|---|---|
| Female | 50.8% | 64.1% | 345 | Increased | **B** |

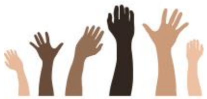

Race

| Race | % in U.S. Population | % in Clinical Trials | Number treated with new drug | Incidence of disease or condition | Grade |
|---|---|---|---|---|---|
| Black or African | 13.4% | 1.6% | 8 | Increased | **F** |
| Asian | 5.9% | 0% | 0 | Decreased | **C** |

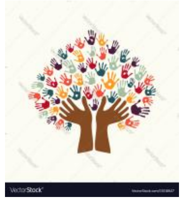

Ethnicity

| Ethnicity | % in U.S. Population | % in Clinical Trials | Number treated with new drug | Incidence of disease or condition | Grade |
|---|---|---|---|---|---|
| Hispanic or Latino | 18.5% | 1.7% | 13 | Decreased | **C** |

**References: Phase 3 trials TG1101-RMS301 and TG1101-RMS302; https://www.accessdata.fda.gov/drugsatfda_docs/nda/2023/761238Orig1s000MedR.pdf pg 105; https://www.nationalmssociety.org/What-is-MS/Who-Gets-MS#:~:text=Research%20has%20demonstrated%20that%20MS,people%20of%20northern%20European%20descent.; https://mymsaa.org/ms-information/overview/who-gets-ms/; 10.1212/WNL.0b013e3182918cc2; https://learningenglish.voanews.com/a/study-majority-of-us-population-now-under-age-40-/5532061.html**

| Drug | |
|---|---|
| Tradename | **Leqembi** |
| Generic Name | **lecanemab-irmb** |
| Company | Eisai Inc. and Biogen |
| Date of FDA Approval | January 6, 2023 |
| Indication | To treat **Alzheimer's disease** |

#### OVERALL GRADE

# C

### CARER Clinical Trial Diversity Scorecard

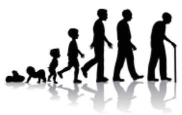

Age

| Age | % in U.S. Population | % in Clinical Trials | Number treated with new drug | Incidence of Disease or Condition | Grade |
|---|---|---|---|---|---|
| >/=65 years old | 16.5% | 80% | 476 | Increased | **A** |

Sex

| Sex | % in U.S. Population | % in Clinical Trials | Number treated with new drug | Incidence of Disease or Condition | Grade |
|---|---|---|---|---|---|
| Female | 50.8% | 50% | 272 | Increased | **C** |

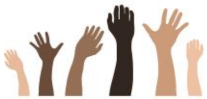

Race

| Race | % in U.S. Population | % in Clinical Trials | Number treated with new drug | Incidence of disease or condition | Grade |
|---|---|---|---|---|---|
| Black or African | 13.4% | 2% | 15 | Increased | **F** |
| Asian | 5.9% | 6% | 36 | Similar | **B** |

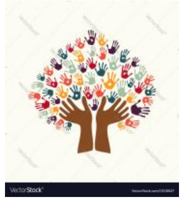

Ethnicity

| Ethnicity | % in U.S. Population | % in Clinical Trials | Number treated with new drug | Incidence of disease or condition | Grade |
|---|---|---|---|---|---|
| Hispanic or Latino | 18.5% | 4% | 26 | Increased | **F** |

**References: Phase 3 trial NCT01767311 Study 201 (BAN2401-G000-201); https://www.brightfocus.org/alzheimers/article/why-does-alzheimers-disease-affect-more-women-men; https://www.accessdata.fda.gov/drugsatfda_docs/nda/2023/761269Orig1s000MedR.pdf pg 266; doi:10.1001/jama.2022.3550**

### CARER Clinical Trial Diversity Scorecard

| Drug | |
|---|---|
| Tradename | **Brenzavvy** |
| Generic Name | **bexagliflozin** |
| Company | TheracosBio, LLC |
| Date of FDA Approval | January 20, 2023 |
| Indication | To improve glycemic control in adults with **type 2 diabetes mellitus** as an adjunct to diet and exercise |

#### OVERALL GRADE

# B

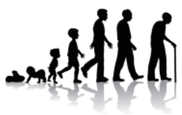

Age

| Age | % in U.S. Population | % in Clinical Trials | Number treated with new drug | Incidence of Disease or Condition | Grade |
|---|---|---|---|---|---|
| >/=65 years old | 16.5% | 43.6% | 871 | Increased | **A** |

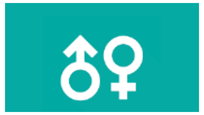

Sex

| Sex | % in U.S. Population | % in Clinical Trials | Number treated with new drug | Incidence of Disease or Condition | Grade |
|---|---|---|---|---|---|
| Female | 50.8% | 38.5% | 751 | Similar | **A** |

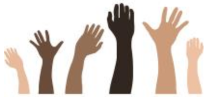

Race

| Race | % in U.S. Population | % in Clinical Trials | Number treated with new drug | Incidence of disease or condition | Grade |
|---|---|---|---|---|---|
| Black or African | 13.4% | 5.7% | 117 | Increased | **D** |
| Asian | 5.9% | 15.9% | 298 | Similar | **A** |

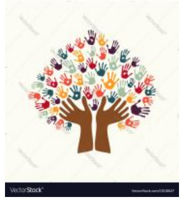

Ethnicity

| Ethnicity | % in U.S. Population | % in Clinical Trials | Number treated with new drug | Incidence of disease or condition | Grade |
|---|---|---|---|---|---|
| Hispanic or Latino | 18.5% | 16.7% | 324 | Increased | **B** |

**References: 6 Phase 3 trials C-419, C-423, C-448, C-450, C-476, C-480 ; doi: 10.2147/IJGM.S226010; https://www.cdc.gov/diabetes/pdfs/library/diabetesreportcard2017**
[https://www.accessdata.fda.gov/drugsatfda_docs/nda/2023/214373Orig1s000IntegratedR.pdf](https://www.accessdata.fda.gov/drugsatfda_docs/nda/2023/214373Orig1s000IntegratedR.pdf) **pg 39**

| Drug | |
|---|---|
| Tradename | **Jaypirca** |
| Generic Name | **pirtobrutinib** |
| Company | Eli Lilly |
| Date of FDA Approval | January 27, 2023 |
| Indication | To treat **relapsed or refractory mantle cell lymphoma** in adults who have had at least two lines of systemic therapy, including a BTK inhibitor |

#### OVERALL GRADE

# B

### CARER Clinical Trial Diversity Scorecard

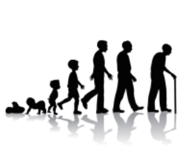

Age

| Age | % in U.S. Population | % in Clinical Trials | Number treated with new drug | Incidence of Disease or Condition | Grade |
|---|---|---|---|---|---|
| >/=65 years old | 16.5% | 77.5% | 93 | Increased | **B** |

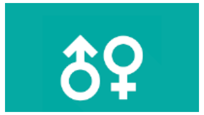

Sex

| Sex | % in U.S. Population | % in Clinical Trials | Number treated with new drug | Incidence of Disease or Condition | Grade |
|---|---|---|---|---|---|
| Female | 50.8% | 20.8% | 25 | Decreased | **C** |

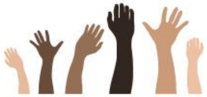

Race

| Race | % in U.S. Population | % in Clinical Trials | Number treated with new drug | Incidence of disease or condition | Grade |
|---|---|---|---|---|---|
| Black or African | 13.4% | 1.7% | 2 | Decreased | **C** |
| Asian | 5.9% | 14.2% | 17 | Decreased | **A** |

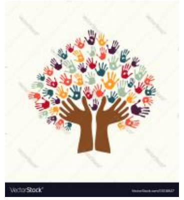

Ethnicity

| Ethnicity | % in U.S. Population | % in Clinical Trials | Number treated with new drug | Incidence of disease or condition | Grade |
|---|---|---|---|---|---|
| Hispanic or Latino | 18.5% | 2.5% | 3 | Decreased | **C** |

**References: Phase 3 trial LOXO-BTK-18001; DOI: 10.1186/1471-2407-14-764;**
**https://www.accessdata.fda.gov/drugsatfda_docs/nda/2023/216059Orig1s000MultidisciplineR.pdf pg 93**

### CARER Clinical Trial Diversity Scorecard

| Drug | |
|---|---|
| Tradename | **Orserdu** |
| Generic Name | **elacestrant** |
| Company | Stemline Therapeutics |
| Date of FDA Approval | January 27, 2023 |
| Indication | To treat estrogen receptor-positive, human epidermal growth factor receptor 2-negative, ESR1-mutated, advanced or **metastatic breast cancer** with disease progression following at least one line of endocrine therapy |

#### OVERALL GRADE

# C

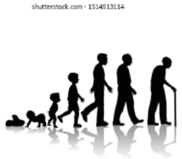


Age

| **Age** | **% in U.S. Population** | **% in Clinical Trials** | **Number treated with new drug** | **Incidence of Disease or Condition** | **Grade** |
|---|---|---|---|---|---|
| >/=65 years old | 16.5% | 55.0% | 104 | Increased | **B** |

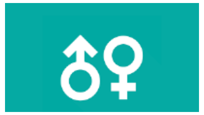

Sex

| **Sex** | **% in U.S. Population** | **% in Clinical Trials** | **Number treated with new drug** | **Incidence of Disease or Condition** | **Grade** |
|---|---|---|---|---|---|
| Female | 50.8% | 98.5% | 233 | Increased | **NA** |

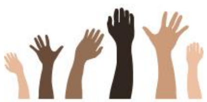

Race

| **Race** | **% in U.S. Population** | **% in Clinical Trials** | **Number treated with new drug** | **Incidence of disease or condition** | **Grade** |
|---|---|---|---|---|---|
| Black or African | 13.4% | 2.7% | 5 | Similar | **D** |
| Asian | 5.9% | 7% | 16 | Decreased | **B** |

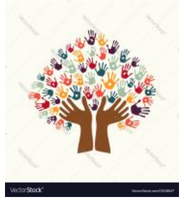

Ethnicity

| **Ethnicity** | **% in U.S. Population** | **% in Clinical Trials** | **Number treated with new drug** | **Incidence of disease or condition** | **Grade** |
|---|---|---|---|---|---|
| Hispanic or Latino | 18.5% | 7.7% | 19 | Decreased | **C** |

**References: Phase 3 trial EMERALD RAD1901-308 ; https://seer.cancer.gov/statfacts/html/breast.html; doi: 10.1200/JCO.22.00338. https://www.cancer.org/content/dam/cancer-org/research/cancer-facts-and-statistics/breast-cancer-facts-and-figures/breast-cancer-facts-and-figures-2019-2020.pdf; https://www.accessdata.fda.gov/drugsatfda_docs/nda/2023/217639Orig1s000MultidisciplineR.pdf pg. 128-129;**

| Drug | |
|---|---|
| Tradename | **Jesduvroq** |
| Generic Name | **daprodustat** |
| Company | GlaxoSmithKline |
| Date of FDA Approval | February 1, 2023 |
| Indication | To treat **anemia caused by chronic kidney disease** for adults on dialysis for at least four months |

#### OVERALL GRADE

# A

### CARER Clinical Trial Diversity Scorecard

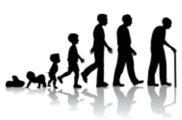

Age

| Age | % in U.S. Population | % in Clinical Trials | Number treated with new drug | Incidence of Disease or Condition | Grade |
|---|---|---|---|---|---|
| >/=65 years old | 16.5% | 32% | >300 | Increased | **A** |

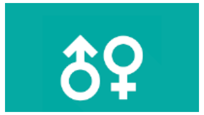

Sex

| Sex | % in U.S. Population | % in Clinical Trials | Number treated with new drug | Incidence of Disease or Condition | Grade |
|---|---|---|---|---|---|
| Female | 50.8% | 43% | >300 | Similar | **A** |

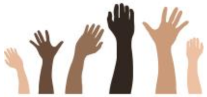

Race

| Race | % in U.S. Population | % in Clinical Trials | Number treated with new drug | Incidence of disease or condition | Grade |
|---|---|---|---|---|---|
| Black or African | 13.4% | 16% | 228 | Increased | **C** |
| Asian | 5.9% | 12% | 176 | Similar | **A** |

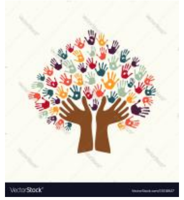

Ethnicity

| Ethnicity | % in U.S. Population | % in Clinical Trials | Number treated with new drug | Incidence of disease or condition | Grade |
|---|---|---|---|---|---|
| Hispanic or Latino | 18.5% | 25% | >300 | Increased | **A** |

**References: Phase 3 trial ASCEND-D; https://www.accessdata.fda.gov/drugsatfda_docs/nda/2023/216951Orig1s000IntegratedR.pdf pg 173; https://www.accessdata.fda.gov/drugsatfda_docs/label/2023/216951s000lbl.pdf; https://www.niddk.nih.gov/health-information/health-statistics/kidney-disease; doi:10.1053/j.semvascsurg.2021.02.010**

| Drug | |
|---|---|
| Tradename | **Lamzede** |
| Generic Name | **velmanase alfa-tycv** |
| Company | CHIESI FARMACEUTICI SPA |
| Date of FDA Approval | February 16, 2023 |
| Indication | To treat **non-central nervous system manifestations of alpha-mannosidosis** |

### CARER Clinical Trial Diversity Scorecard

**OVERALL GRADE**

**D**

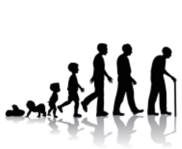

**Age**

| Age | % in U.S. Population | % in Clinical Trials | Number treated with new drug | Incidence of Disease or Condition | Grade |
|---|---|---|---|---|---|
| >/=65 years old | 16.5% | 0% | 0 | Decreased | **NA** |

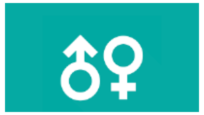

**Sex**

| Sex | % in U.S. Population | % in Clinical Trials | Number treated with new drug | Incidence of Disease or Condition | Grade |
|---|---|---|---|---|---|
| Female | 50.8% | 44% | 6 | Similar | **C** |

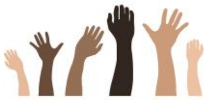

**Race**

| Race | % in U.S. Population | % in Clinical Trials | Number treated with new drug | Incidence of disease or condition | Grade |
|---|---|---|---|---|---|
| Black or African | 13.4% | 0% | 0 | Similar | **D** |
| Asian | 5.9% | 0% | 0 | Similar | **D** |

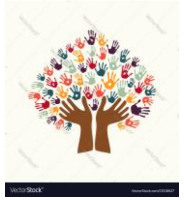

**Ethnicity**

| Ethnicity | % in U.S. Population | % in Clinical Trials | Number treated with new drug | Incidence of disease or condition | Grade |
|---|---|---|---|---|---|
| Hispanic or Latino | 18.5% | NR | NR | Similar | **D** |

**References: Phase 3 trial rhLAMAN-05 https://rarediseases.org/rare-diseases/alpha-mannosidosis/#affected**
https://doi.org/10.1186/s13023-022-02422-6; https://www.accessdata.fda.gov/drugsatfda_docs/nda/2023/761278Orig1s000IntegratedR.pdf
**pg28**

### Geographic representations from US or North America in Phase 3 Pivotal trials

- Mounjaro 22.0% from the US
- Voquezna 42.7% from US
- Enjaymo- 12.5% from US
- Camzyos  43.0% from US
- Odpaulag- 11.1% from USA/Canada
- Ztalymy 41.6% from US
- Vivjoa 53% from US
- Quviviq 33% from US
- Spevigo 6% from US
- Sotyku 32% from US
- Terlivaz 89%
- Relyvrio 100%
- Lytgobi 45.6% from North America
- Rolvedon 81.5%
- Elucirem 25.8% from North America
- Tecvayli 33.1% US
- Elahere 22% US

Tzield 95% US

Imjudo 7.7%

Leqembi 80% from US

Briumvi 7.7% from US

NexoBrid 29.6% from US

Brenzavvy 36.1%

Lamzede 0%

Jesduvroq 29%

Kimmtrak 31.7% in US

Vtama 75.7%

Amvuttra 20.1%

Xenpozyme 13.9%

Krazati 100%

Lunsumio 44.4%

Xenoview 100%
